# Multi-ancestry genome-wide association study of endometriosis and its clinical manifestations in ∼1.4 million women: translating gene discovery into pathogenic mechanisms and therapeutic targets

**DOI:** 10.1101/2025.09.03.25335012

**Authors:** Dora Koller, Jun He, Solveig Løkhammer, Selena Aranda, Dan Qiu, David Davtian, Qianyu Chen, Ziang Xu, Zhongzheng Mao, Eleni Friligkou, Sefayet Karaca, Bru Cormand, Idhaliz Flores, Signe Altmäe, Marina Mitjans, Brenda Cabrera-Mendoza, Renato Polimanti

## Abstract

We conducted a multi-ancestry genome-wide association study of endometriosis and adenomyosis in ∼1.4 million women, including 105,869 cases, aiming to expand endometriosis loci discovery across ancestries, dissect symptom-specific effects, and integrate multi-omic data. We identified 80 genome-wide significant associations, 37 of which are novel, including five loci that are the first ever variants reported for adenomyosis. Fine-mapping and colocalization analyses uncovered causal loci for over 50 endometriosis-related associations. Multi-omics integration revealed that genetic variation influences endometriosis risk through transcriptomic, epigenetic, and proteomic regulation across multiple tissues, converging on pathways involved in immune regulation, tissue remodeling, and cell differentiation. Drug-repurposing analyses highlighted potential therapeutic interventions currently used for breast cancer and preterm birth prevention. Endometriosis polygenic risk interacted with abdominal pain, anxiety, migraine, and nausea. This study advances the understanding of genetic risk factors for endometriosis and provides molecular support for several hypotheses on the disease’s pathogenesis.

## INTRODUCTION

Endometriosis is a chronic, systemic inflammatory disease characterized by the presence of endometrial-like tissue outside the uterine cavity ^1^. The disease, which affects 10% of reproductive-age female participants, carries a significant public health burden due to a debilitating, multi-system symptomatic profile that impacts both physical and mental health and severely reduces quality of life ^2^. While endometriosis is a complex disease, it has a substantial genetic component, with a twin-based heritability estimated at 50% ^3^ and a single nucleotide polymorphism (SNP)-based heritability (SNP-*h*^2^) of 8% ^4,5^. Genome-wide association studies (GWAS) have been instrumental in dissecting the biology of endometriosis, identifying several risk loci. These discoveries have provided crucial insights into the molecular pathways involved in disease pathogenesis, implicating genes related to hormonal regulation, immunity, and cellular adhesion ^4,6,7^. The two most recent GWASs of endometriosis, comprising 928,413 participants (including 44,125 cases; 30% non-European ancestry) ^7^ and 762,600 participants of European descent (including 60,674 cases) ^4^, identified up to 45 loci, expanding the understanding of the disease pathogenesis.

Nevertheless, the complex genetic architecture of endometriosis remains to be fully characterized, especially related to the range of symptoms reported by affected women. Previous efforts to address endometriosis heterogeneity have included stratification by surgically defined subtypes, pain-related phenotypes, and the presence of comorbid conditions such as adenomyosis (a disease where endometrial tissue grows into the muscular wall of the uterus ^8^) and infertility ^4^. However, there is no international consensus on how to define or classify endometriosis. Existing surgical classification and staging systems show little or no correlation with patient outcomes, including pain severity and other symptoms ^9^. Current expert consensus suggests that evaluating disease localization and associated symptoms may be more informative than relying solely on staging systems ^9^.

In this study, we conducted a large-scale cross-ancestry GWAS including 105,869 cases and 1,282,731 controls to substantially expand the map of genetic risk factors for endometriosis and to further dissect its clinical and genetic heterogeneity, and report the first genome-wide significant loci for adenomyosis. Moving beyond traditional classifications, we performed novel genetic analyses on a wide range of clinical symptoms and explored the genetic overlap with key comorbidities related to women’s health, allowing for an in-depth investigation of pleiotropy and potential causal relationships. While ancestry-specific analyses have been previously conducted, our study is the first to implement a cross-ancestry polygenic risk score (PRS) framework, including individuals of six ancestry groups (African, AFR; Admixed American, AMR; Central/South Asian, CSA; East Asian, EAS; European, EUR; Middle Eastern, MID) to assess predictive performance and genetic transferability across global populations. To translate genetic associations into biological function, we performed the first comprehensive functional analyses for different definitions of endometriosis and adenomyosis. Overall, this work refines our understanding of the genetic architecture of endometriosis, providing new insights into the biology underlying its specific clinical features, and contributes to demonstrate the potential usefulness of genetic information to develop precision medicine approaches.

## RESULTS

### Genetic Discovery, Heritability and Genetic Correlation across Cohorts

We combined GWAS of endometriosis from eight cohorts consisting of six ancestries (**Supplemental Table 1**). In the EUR endometriosis cohort, statistically significant SNP-*h^2^* was observed in all cohorts except for MVP (z=1.10), with z-scores ranging from 5.88 (AoU) to 13.62 (23andMe, Inc.). The genetic correlation (rg) among endometriosis cohorts ranged from 0.72 (p = 4.85 × 10^−13^) between AoU and EstBB to 1.05 (p < 2.2 × 10^-308^) between FinnGen and EstBB, with a median rg estimate of 0.87. Adenomyosis had significant SNP-*h^2^* only in the FinnGen cohort (z=5.88), and endometriosis without adenomyosis had significant h2 in both AoU and UKB (z=4.07 and z=4.48, respectively). Significant SNP-*h^2^* was also observed for both clinical (ICD-10 N80 or SNOMED-129103003) and self-reported endometriosis definitions in both cohorts (clinical z_AoU_=2.76 and z_UKB_=6.12; self-reported z_AoU_=3.43 and z_UKB_=3.07) and they were all genetically correlated (rg ranging from 0.92 (p=1.4 × 10^−7^) between AoU clinical and self-reported definitions to 1.03 (p=7 × 10^−4^) between AoU clinical and UKB clinical definition (**Supplemental Table 2)**. EUR-specific and cross-ancestry meta-analyzed data all had significant SNP-*h^2^*with z-scores ranging from 5.66 (EUR without adenomyosis) to 16.41 (EUR endometriosis) with no evidence of inflation (linkage disequilibrium score regression, LDSC intercept<1.04; **Supplemental Table 3**). However, deflation was observed in the EUR-specific GWAS meta-analysis due to the sample-overlap correction applied (see Methods). We did not observe statistically significant SNP-*h^2^* among non-EUR populations owing to their limited sample size, except for AMR (z=2.15). All cross-ancestry and EUR meta-analyses of endometriosis were genetically correlated, ranging from 0.90 (p<2.2×10^-308^, between EUR combined endometriosis and EUR clinical endometriosis) to 1 (p<2.2×10^-308^, between cross-ancestry endometriosis and EUR endometriosis) (**Supplemental Table 3**). The genetic correlation of adenomyosis ranged from 0.79 (p=9.76×10^-67^) with EUR combined endometriosis to 0.89 (p=2.13×10^-96^) with endometriosis clinical definition.

### Gene Discovery

We identified 80 LD-independent (r^2^<0.1) variants with genome-wide significant (GWS) association (p<5×10^−8^) with endometriosis combined definition (i.e., combining clinical and self-reported information) in the cross-ancestry meta-analysis (**Fig. 1** and **Supplemental Table 4**). Of these, 37 were novel (LD-independent from the previous GWAS findings). Among the new associations, the most significant was rs10514243 (β=-0.01, p=5.9×10^−13^), located near *CTD-2218K11.2*, a predicted long non-coding RNA locus, ∼280 kb downstream from the *XRCC4* gene and ∼250 kb upstream from the *RPS23* and *ATG10* genes. When stratifying the European sample by data source, this locus and most other cross-ancestry associations showed stronger statistical significance in the endometriosis clinical definition sample compared with other endometriosis definitions (EUR clinical endometriosis: β=-0.02, p=2×10^−9^, EUR self-reported endometriosis: β=-0.02, p=1.9×10^−6^; EUR adenomyosis: β=-0.03, p=7.4×10^−4^; **Supplemental Table 4**). The previously reported *SYNE1* rs58415480 locus was significant for all endometriosis definitions and adenomyosis in EUR (cross-ancestry endometriosis: β=-0.04, p=9.7×10^−62^, EUR combined endometriosis: β=-0.03, p=6×10^−57^, EUR clinical endometriosis: β=-0.05, p=8.1×10^−58^, EUR self-reported endometriosis: β=-0.05, p=1.7×10^−20^, adenomyosis: β=-0.07, p=6.2 × 10^−13^, and endometriosis without adenomyosis: β=-0.09, p=1.5×10^−9^). Of the 80 loci, *WNT4* rs55938609 reached genome-wide significance (β=0.1, p=2.5×10^−13^) in EAS, which was also genome-wide significant for all EUR definitions, except for adenomyosis (**Supplemental Table 4**).

**Figure 1.**
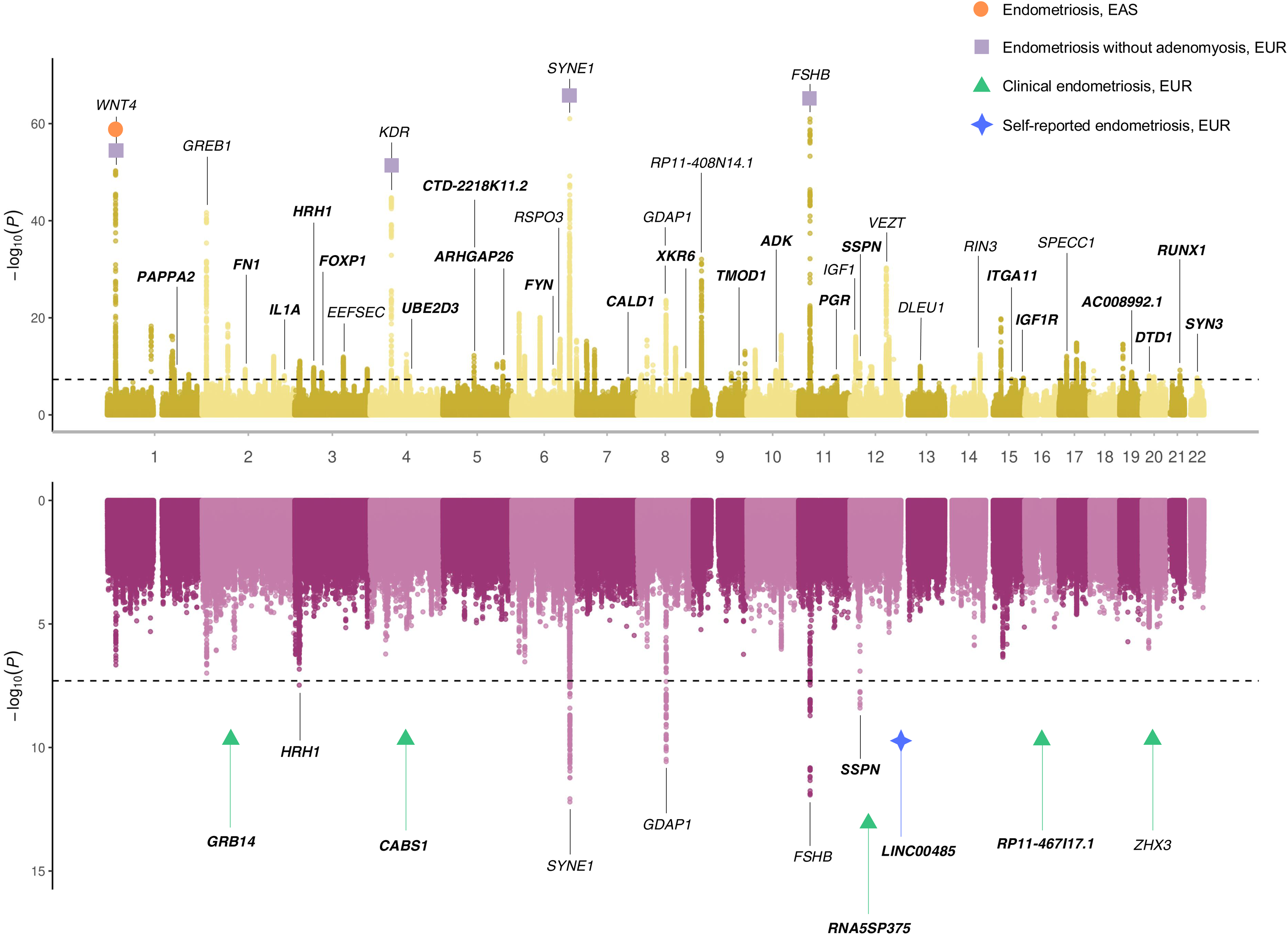
Genome-wide association study (GWAS) of endometriosis (top) and adenomyosis (bottom). Manhattan plots related to the cross-ancestry GWAS meta-analysis of endometriosis combined definition (n_cases_=105,869 and n_controls_=1,282,731) and adenomyosis (n_cases_=8,763 and n_controls_=415,718). Loci highlighted in bold represent novel hits compared to previous endometriosis GWASs. Loci reaching significance in East Asian and European descent are highlighted with a circle and a square, respectively. Each point represents the p-value from a two-sided test of association between individual genetic variants and the phenotype, ordered by genomic position on the x axis and shown as −log10(p-value) on the y axis.

We identified 63 genome-wide significant loci for the EUR endometriosis combined definition, one for EAS, 44 for EUR clinical endometriosis, 30 for EUR self-reported endometriosis, five for EUR adenomyosis, and five for EUR endometriosis without adenomyosis (**Supplemental Table 4**). We did not identify any genome-wide significant SNPs for the other population groups. Clinical endometriosis revealed four novel loci (most significant locus: rs12444457 β=0.02, p=1.1×10^−8^, located near *RP11-467I17.1*, approximately 160 kb downstream of *WWOX* and ∼230 kb upstream of *MAF*) that were not genome-wide significant in the other endometriosis definitions investigated in the present study (**Supplemental Table 4**). Additionally, there was one locus specific to self-reported endometriosis (rs10860902 (β=0.02, p=1×10^−8^), located near *LINC00485*, ∼135 kb upstream of *IGF1* and ∼145 kb downstream of *PAH*). Two novel endometriosis-associated loci, *HRH1* rs184700 and *SSPN* rs7974900 were also genome-wide significant for adenomyosis (β=0.06, p=3.3×10^−8^ and β=0.05, p=6.3×10^−9^, respectively), but they did not reach significance for endometriosis without adenomyosis.

Considering LD independent variants (r^2^<0.1) from the index SNPs in the primary GWAS, the conditional analysis identified five additional loci for the endometriosis combined phenotype and eight additional loci for the EUR clinical phenotype (p_J_<5 × 10^-8^; **Supplemental Table 5**). Among these loci, two variants mapped to the *ESR1* gene: rs74713348 (cross-ancestry combined definition p_J_=6.97×10^-15^; EUR combined definition p_J_=8.43×10^-14^; EUR clinical definition: p_J_=5.49×10^-21^) and rs111930025 (cross-ancestry combined definition p_J_=7.9×10^-10^; EUR combined definition p_J_=4.84×10^-9^; EUR clinical definition: p_J_=7.7×10^-10^).

### Multi-Omics Characterization

To prioritize plausible causal variants within the LD-independent genome-wide significant loci, we conducted a fine-mapping analysis using PAINTOR ^10^, incorporating Bonferroni-significant functional annotations (**Supplemental Table 6**). Following PAINTOR recommendations ^10^, we included only minimally correlated, endometriosis-relevant annotations. For the EUR combined and cross-ancestry datasets, selected annotations included histone H3 trimethylated at lysine 4 (H3K4me3) and histone H3 acetylated at lysine 27 (H3K27ac) in uterine and ovarian tissues, super-enhancers in ovarian tissue, and differentially expressed enhancers in uterine tissue. A similar set was used for the EUR-clinical cohort, with annotations for H3K27ac, super-enhancers in ovary tissue, and differentially expressed enhancers in uterus tissue. For the EAS dataset, no annotations met the Bonferroni threshold, and fine-mapping was therefore performed without functional priors. Across ancestry-specific and cross-ancestry analyses of endometriosis definitions, we identified putative causal evidence (posterior probability, PP>0.8) for 21 variants (**Supplemental Table 7**). Among these, rs28858973 and rs62532633 showed causal evidence in both EUR-specific analysis (PP=0.91 and 1, respectively) and cross-ancestry analysis (PP=0.95 and 1, respectively). Cross-ancestry analysis also identified rs7926666, rs17428012, and rs1964592 (PP=1, 0.84, and 0.99, respectively), which showed suggestive evidence in EUR (PP=0.79, 0.59, and 0.59). Among variants identified as causal considering EUR endometriosis clinical definition, rs12108558 showed suggestive causal evidence in EUR endometriosis combined definition (PP_EUR-clinical_=0.87; PP_EUR-combined_=0.70), while rs117914842 showed suggestive evidence in the cross-ancestry analysis (PP_EUR-clinical_ =0.9; PP_cross-ancestry_=0.60). In the EAS analysis, convergent suggestive evidence in EAS, EUR, and cross-ancestry combined definitions were observed for rs2501284 (PP=0.59, 0.6, and 0.59, respectively). In total, suggestive causal evidence (PP>0.5) was identified for 63 additional variants (**Supplemental Table 7**).

We identified 29 colocalization signals (posterior probability of full colocalization, PPFC>0.7) related to 13 SNPs (posterior probability for an individual SNP, p_snp_>0.5) showing shared causal effects on endometriosis and tissue-specific transcriptomic regulation (**Supplemental Table 8**). For the EUR clinical endometriosis definition, rs13100173 showed causal effect on endometriosis and *HYAL3* transcription regulation in vagina (PPFC=0.71, p_snp_=0.75), uterus (PPFC=0.82, p_snp_=0.72), ovary (PPFC=0.81, p_snp_=1), breast mammary tissue (PPFC=0.81, p_snp_=0.85), sigmoid colon (PPFC=0.81, p_snp_=0.94), and transverse colon (PPFC=0.81, p_snp_=0.93). Broad cross-tissue colocalization was also observed for rs61926299 in the cross-ancestry endometriosis combined definition and *ATF1* transcription regulation in ovary (PPFC=0.71, p_snp_=0.56), breast mammary tissue (PPFC=0.95, p_snp_=0.85), sigmoid colon (PPFC=0.92, p_snp_=0.58), and transverse colon (PPFC=0.81, p_snp_=0.91); and *LIMA1* in transverse colon (PPFC=0.93, psnp=0.69). Convergent colocalization between endometriosis combined and clinical definitions was observed for rs9739834 with respect to *VEZT* transcriptomic regulation in sigmoid colon (combined definition: PPFC=0.73, p_snp_=1; clinical definition: PPFC=0.86, p_snp_=1). While the majority of the colocalization signals were observed with respect to EUR and cross-ancestry associations, we also identified a colocalization for rs2388493 between EAS endometriosis combined definitions and *BAALC* transcription regulation in sigmoid colon (PPFC=0.81, p_snp_=0.56)

The transcriptome-wide association study (TWAS) combining tissues implicated in the pathogenesis of endometriosis and its symptoms (i.e., uterus, vagina, ovary, breast mammary tissue, whole blood, colon sigmoid, and colon transverse) identified 41 genes after Bonferroni multiple testing correction (p<3.72×10^-6^; **Supplemental Table 9** and **Fig. 2**). Top findings included a negative association of *ARL14EP* expression with endometriosis risk (p_multi-tissue_=1.36×10^-40^; best tissue: breast mammary tissue), a positive association of *CDC42* expression (p_multi-tissue_=3.76×10^-20^; best tissue: colon sigmoid), and *CCDC170* (p_multi-tissue_=4.33×10^-17^; best tissue: whole blood). Increased predicted expression of *RBM6* was associated with a reduced risk of endometriosis in the multi-tissue analysis (p=1.79×10^-6^), the primary tissues affected by the disease (p_uterus_=2.05×10^-7^; p_ovary_=5X10^-7^; p_vagina_=1.63×10^-6^), and whole blood (p=3.17×10^-6^). Two additional genes identified in the multi-tissue TWAS showed positive associations with endometriosis (*GMPPB* p_multi-tissue_=5.59×10^-8^, p_uterus_=1.92×10^-7^; *CRISPLD1* p_multi-tissue_=9.26×10^-8^, p_uterus_=1.55×10^-9^). Among multi-tissue TWAS significant results, ovary- and vagina-specific positive associations with endometriosis were present for *WASHC3* (p_multi-tissue_=2.11×10^-9^; p_ovary_=1.49×10^-8^; p_vagina_=6.7×10^-8^) and *DTD1* (p_multi-tissue_=4.1×10^-7^; p_ovary_=1.47×10^-8^; p_vagina_=7.77×10^-9^). While they did not reach Bonferroni correction in the multi-tissue TWAS, we observed positive endometriosis associations of *CISD2* (z=4.78, p=1.75×10^-6^) in the uterus-specific analysis, and lower expression of *EIF2AK4* (z=-4.99, p=6.05×10^-7^) and *MSRA* (z=-4.68, p=2.81×10^-6^) in the vagina-specific analysis.

**Figure 2.**
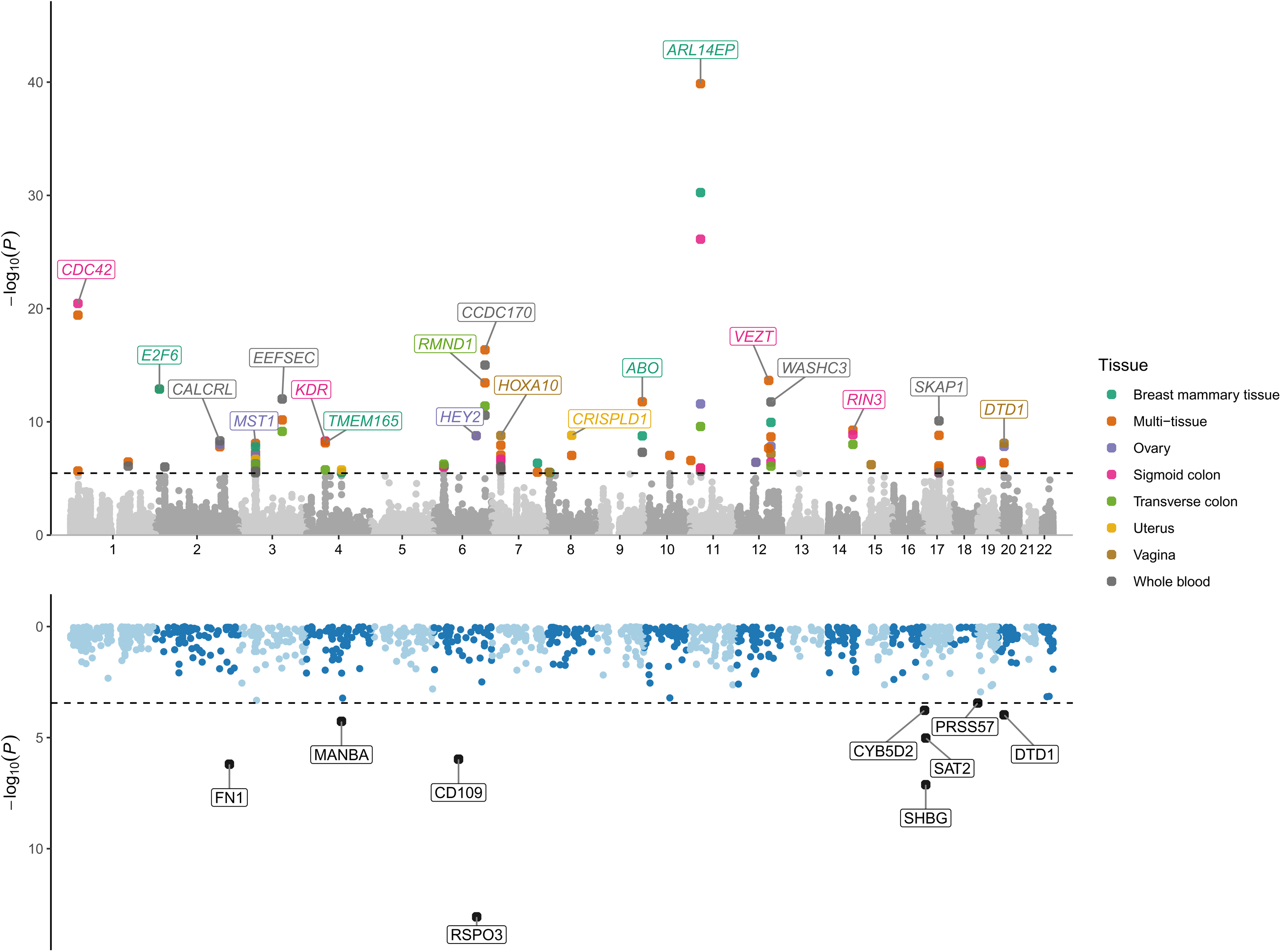
Transcriptome-wide (top) and proteome-wide (bottom) associations of endometriosis combined definition. Transcriptome-wide associations are those obtained from the multiple-tissue analysis. Each point represents the p-value from a two-sided z-test of association between the phenotype and the cis-genetically regulated expression level of a gene or plasma protein, ordered by genomic position on the x axis and shown as −log10(p-value) on the y axis. The black horizontal dashed lines indicate the significance thresholds after multiple testing correction for the total number of models (p<3.4×10^-6^ for TWAS and p<3.6×10^-4^ for PWAS). In the top panel, the colors of the points represent different tissues, and gene names are colored based on the most significant tissue.

We identified nine proteins whose genetically predicted plasma levels were significantly associated with endometriosis risk (FDR q<0.05; **Supplemental Table 10** and **Fig. 2**). The strongest proteome-wise association was observed for *RSPO3*, which showed a positive association with endometriosis in two distinct pQTL models (z=7.46 and 7.38; p=8.52×10^-14^ and 1.54×10^-13^, respectively). Other FDR-significant positive proteome-wide associations included *SHBG* (z=5.38, p=7.61×10^-8^), *SAT2* (z=4.43, p=9.56×10^-6^), and *PRSS57* (z=3.57, p=3.61×10⁻^4^). Conversely, *FN1* was the most significant negative proteome-wide association (z=–4.98, p=6.36×10^-7^). Other negative proteome-wide effects included *CD109* (z=–4.88, p=1.07×10^-6^), *MANBA* (z=-4.04, p=5.42×10^-5^), *DTD1* (z=–3.88, p=1.06×10^-4^), and *CYB5D2* (z=–3.76, p=1.69×10⁻^4^).

In EUR, blood-based SMR meta-analysis identified 305 CpG sites associated with endometriosis (FDR q<0.05, HEIDI p>0.05), including 242 for the combined definition (**Supplemental Table 11**), 62 for the clinical definition (**Supplemental Table 12**), and 49 for the self-reported definition (**Supplemental Table 13**). Five CpG sites within three loci reached FDR-significant associations without evidence of heterogeneity across the three endometriosis definitions investigated: cg10740902 and cg07098391 mapping on *KDR* gene; cg09995736 near *VEZT* gene; cg08907436 and cg17706972 mapping on *ESR1* gene. The latter associations are located within a genomic region that includes 12 additional FDR-significant CpG sites (top-result: cg23164938 p_combined_=5.49×10^-11^). In the MHC region (chromosome 6: 29,600,203-31,129,828), there were 29 convergent FDR-significant effects (top-CpG association: cg14574237 p=6.63×10^-7^; 14 related to endometriosis combined definition and 15 to self-reported endometriosis with five CpG sites shared between the two). Another locus with convergent SMR results was observed on chromosome 12 (25,886,475-27,260,441) mapping several non-coding genes and three protein-coding genes (*CBX3*, *SNX10*, *HNRNPA2B1*, and *NFE2L3*), where 23 CpG associations related to the three endometriosis definitions (top-result: cg12942962 p_combined_=1.25×10^-8^). Because of the limited power of the adenomyosis SMR analysis, we identified only three FDR-significant CpG associations that were also identified in the endometriosis analyses: cg11466449 (p_adenomyosis_=1.37×10^-6^) mapping to *ENSG00000289405* RNA gene and cg12868707 (p_adenomyosis_=1.83×10^-8^) and cg22105613 (p_adenomyosis_=7.23×10^-7^) both mapping to *GDAP1* gene (**Supplemental Table 14**). The latter CpG site was also identified in combined (p=4.88×10^-10^) and clinical (p=9.63×10^-7^) endometriosis. While we did not identify FDR significant results in the EAS blood-based SMR analysis, cross-ancestry nominally significant replication of EUR findings was observed for eight CpG sites (combined definition: cg00814883, cg02971253, cg02060096, cg07227024, cg09678539, clinical definition: cg14396995, cg13172367, and self-reported definition: cg16650630).

In the endometrium-specific SMR, we identified 57 CpG sites associated with endometriosis (FDR q<0.05; **Supplemental Table 15**). In the endometriosis combined definition, the top finding was cg03519931 mapping to *WNT4* gene (p=1.65×10^-14^). This CpG site was also FDR significant for the endometriosis clinical definition (p=7.22×10^-10^) and adenomyosis (p=1.43×10^-5^). Also, the other two CpG sites identified in the adenomyosis SMR analysis (cg13258528 p=7.87×10^-6^, cg10414477 p=1.96×10^-5^) were also significantly associated with endometriosis (p_self-reported_=2.98×10^-10^ and 1.03×10^-5^, respectively). Although there was consistency across endometriosis definitions and between endometriosis and adenomyosis for most associations, we also observed 20 CpG sites that reached FDR significance for endometriosis clinical definition, but were null for self-reported endometriosis (p>0.05; **Supplemental Table 15**). The top clinical vs. self-reported difference was cg05678033 (p=1.31×10^-6^ vs. 0.239). Conversely, only two CpG sites were FDR significant with respect to self-reported endometriosis but null with respect to endometriosis clinical definition (cg06488957 mapping on *RAI1* gene and cg22228116 mapping on *HLCS* gene; **Supplemental Table 15**) With respect to cross-tissue convergence, cg24126931 mapping on *HCG2P7* gene was FDR significant in blood (p_combined_=5.69×10^-5^; p_self-reported_=4.03×10^-5^) and endometrium (p_combined_=6.41×10^-5^; p_self-reported_=4.73×10^-6^).

### Drug repositioning

We applied DRUGSETS ^11^, a genetically informed drug repositioning framework, to identify therapeutic options for endometriosis. Considering EUR gene-based results related to endometriosis combined definition, we found two Bonferroni-significant drugs, neratinib and toremifene (p<6.8×10^-5^; **Supplemental Table 16**). The cross-ancestry analysis identified three additional Bonferroni-significant (Bonferroni) candidates: 17-hydroxyprogesterone-caproate, abemaciclib, and norgestimate (p<6.8×10^-5^; **Supplemental Table 17**).

### Gene Ontology Enrichment

After applying Bonferroni correction (p<6.31×10⁻⁷), we identified a total of 1,727 GO terms significantly enriched for endometriosis considering the EUR GWAS combined definition (**Supplemental Table 18**). Top results included “negative regulation of response to stimulus” (GO:0048585, p=6.1×10⁻^85^), “regulation of immune system process” (GO:0002682, p=2.12×10⁻^74^), and “tissue development” (GO:0009888, p=1.41×10⁻^70^). Through REVIGO ^12^, we identified 1,194 non-redundant GO terms enriched for EUR endometriosis combined definition. Among these, 70% were associated with 118 functional themes and the three largest of them were “regulation of immune response” (GO-term N=43), “regulation of transport” (GO-term N=42), and “regulation of cell differentiation” (GO-term N=32) (**Supplemental Table 19**). For adenomyosis, we identified 832 Bonferroni-significant GO enrichments (**Supplemental Table 18**). Among these, 817 GOs (98%) were Bonferroni significant also for endometriosis. However, the GO terms shared between endometriosis and adenomyosis, we identified five gene sets that were statistically more enriched for endometriosis than for adenomyosis (**Supplemental Table 18**; p<0.001): “regulation of cell population proliferation” (GO:0042127; β_endometriosis_=1.54, β_adenomyosis_=1.04, p=1.2×10⁻^4^), “epithelial cell proliferation” (GO:0050673; β_endometriosis_=2.29, β_adenomyosis_=1.14, difference-p=1.5×10⁻^4^), “epithelial cell differentiation” (GO:0030855; β_endometriosis_=1.83, β_adenomyosis_=1.15, difference-p=3.2×10⁻^4^), “positive regulation of catalytic activity” (GO:0043085; β_endometriosis_=1.58, β_adenomyosis_=1.04, difference-p=3.79×10⁻^4^), and “positive regulation of cell population proliferation” (GO:0008284; β_endometriosis_=1.66, β_adenomyosis_=1.05, p=4.61×10⁻^4^).

### Polygenic Risk Scoring

EUR endometriosis-PRS derived from non-overlapping cohorts was significantly associated with AoU-endometriosis (odds ratio, OR=1.38, 95%CI=1.35-1.41) and UKB-endometriosis (OR=1.3, 95%CI=1.27-1.34) in EUR, and also with endometriosis across different population groups in AoU. Compared to individuals in the lowest PRS quartile, those in the highest quartile showed increased odds of endometriosis in EUR-AoU (OR=2.19, 95%CI=2.07– 2.32), AMR-AoU (OR=1.77, 95%CI=1.54–2.04), EAS-AoU (OR=2.75, 95%CI=1.81–4.27), AFR-AoU (OR=1.44, 95%CI=1.23–1.69), SAS-AoU (OR=1.92, 95%CI=1.06–3.55), and MID-AoU (OR=3.68, 95%CI=1.27–12.20) (**Fig. 3** and **Supplemental Table 20**). In addition to being affected by the sample size of the target samples, the magnitude of cross-ancestry PRS effects generally reflected the genetic distance between EUR and the other population groups.

**Figure 3.**
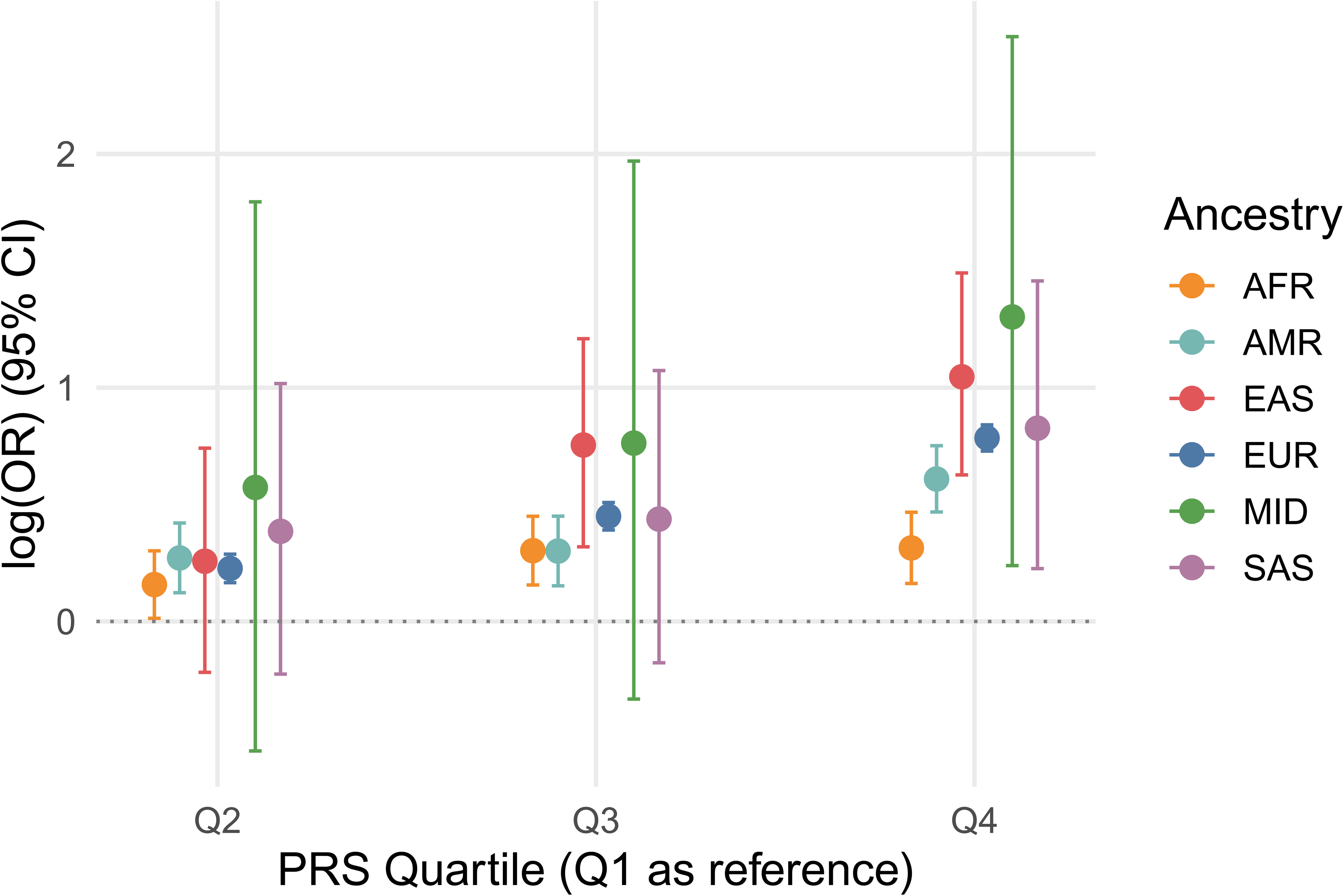
Within-ancestry and cross-ancestry PRS associations of endometriosis. logORs and corresponding 95% confidence intervals are reported. In the within-ancestry analysis, the EUR-endometriosis PRS (excluding EUR-AoU from the training sample, N_eff_=322,319) is tested against the EUR-AoU sample (N_eff_=42,118). In the cross-ancestry analysis, EUR-endometriosis PRS (with EUR-AoU in the training sample, N_eff_=364,494) is tested against AoU samples of AFR (N_eff_=6,802), AMR (N_eff_=6,617), EAS (N_eff_=832), MID (N_eff_=129), and SAS (N_eff_=409) descent.

We performed interaction analyses between endometriosis-related symptoms and the EUR endometriosis-PRS across two independent cohorts: AoU and UKB (**Supplemental Table 21**). Specifically, we tested whether the presence of individual symptoms modifies the effect of the endometriosis PRS on endometriosis risk. Results surviving FDR multiple comparisons included PRS×abdominal pain (OR=0.94, 95%CI=0.91-0.98), PRS×anxiety (OR=0.93, 95%CI=0.89-0.96), PRS×migraine (OR=0.94, 95%CI=0.91-0.98), PRS×nausea (OR=0.90, 95%CI=0.87-0.95), PRS×comorbidity burden (OR=0.99, 95%CI=0.98-0.99) in AoU (**Fig. 4** and **Supplemental Table 22**). All showed negative interaction, so attenuation of the PRS effect in individuals reporting the symptom. While the AoU interactions were not statistically replicated in UKB, there was no AoU-UKB difference (difference-p<0.05; **Supplemental Table 22**) in the interaction statistics for abdominal pain, anxiety, and comorbidity burden.

**Figure 4.**
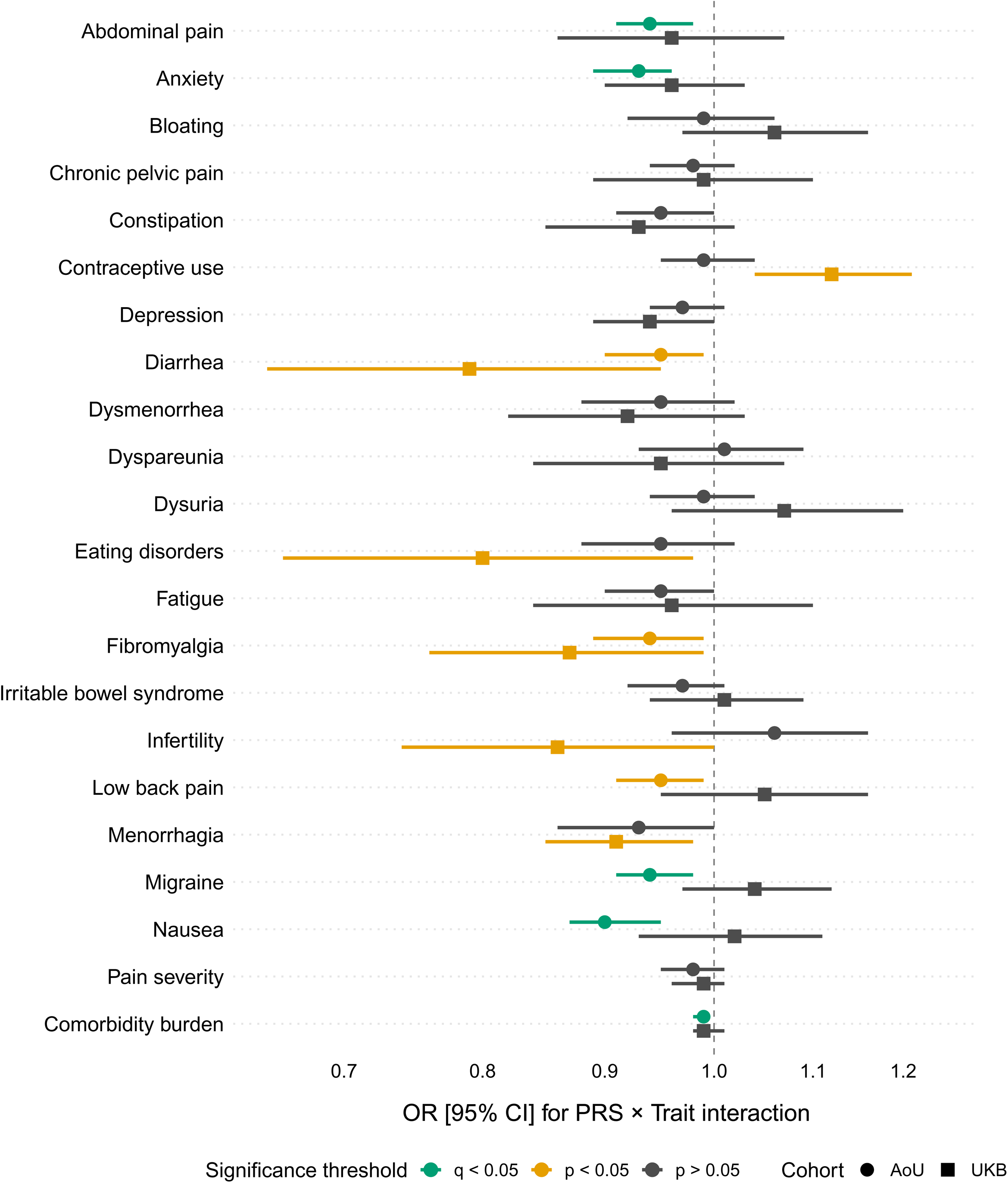
Interaction effects between endometriosis PRS and endometriosis symptoms and comorbidities in AoU and UKB. Forest plot illustrating the OR and 95% CI for the interaction between endometriosis PRS and its various symptoms and comorbidities in two independent cohorts: AoU (circles) and UKB (squares). Each row represents a different symptom or comorbidity, with the x-axis showing the OR for the PRS×trait interaction on endometriosis risk. Green points indicate results significant after false discovery rate (FDR)-correction (q<0.05). Yellow points indicate nominal significance (p<0.05), and grey points indicate non-significant results (p>0.05).

By contrast, nominally significant AoU-UKB differences were observed for migraine (p_difference_=0.02) and nausea (p_difference_=0.03).

### Pleiotropy of Endometriosis with Comorbid Conditions

We investigated genetic correlations between endometriosis and 35 related traits encompassing gynecological/reproductive, psychiatric, pain-related, metabolic, gastrointestinal, cancer, and systemic categories (**Supplemental Table 23**). Considering heritable phenotypes (SNP-*h^2^* p<0.05; **Supplemental Table 24**), endometriosis combined definition showed 23 significant genetic correlations (FDR q< 0.05), predominantly involving gynecological and reproductive traits such as pain and other conditions associated with female genital organs and menstrual cycle (rg=0.80, p=9.14×10^-58^), excessive, frequent, and irregular menstruation (rg=0.75, p=4.5×10^-157^), ovarian cysts (rg=0.78, p=2×10^-53^), female infertility (rg=0.64, p=4.9×10^-49^), leiomyoma of the uterus (rg=0.44, p=6.7×10^-41^), polyp of female genital tract (rg=0.53, p=2.9×10^-18^), inflammatory disease of the uterus (rg=0.61, p=1.2×10^-12^), hemorrhage in early pregnancy (rg=0.40, p=1.3×10^-6^). Significant genetic correlation was also found with major depressive disorder (MDD) (rg=0.37, p=1.66×10^-72^), anxiety (rg=0.40, p=1.09×10^-47^) (**Fig. 5** and **Supplemental Table 24**).

**Figure 5.**
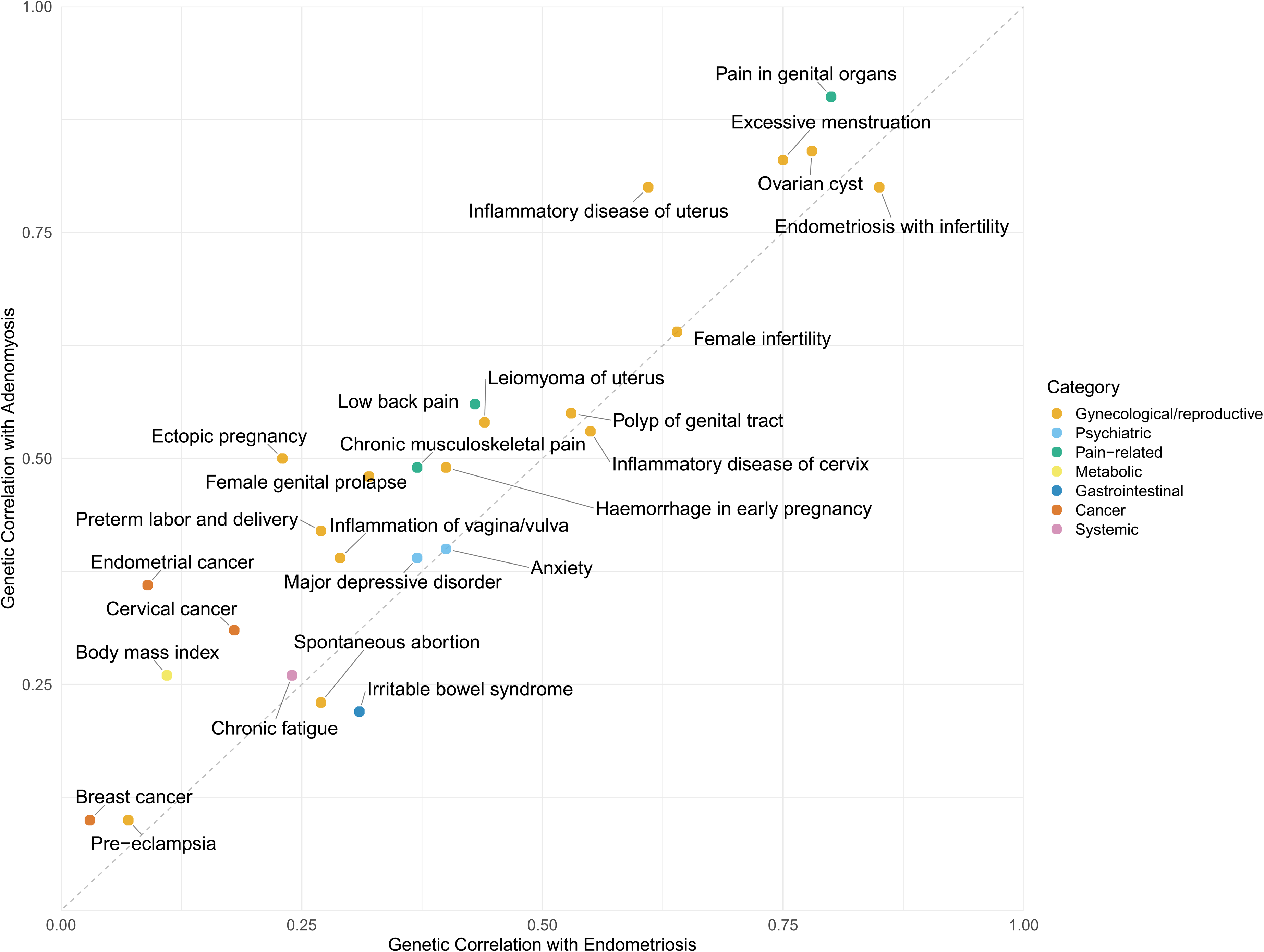
Genetic correlations of endometriosis with gynecological/reproductive, psychiatric, pain-related, metabolic, gastrointestinal, cancer, and systemic traits. Genetic correlations with endometriosis (combined definition) and the selected traits are shown on the x axis, while genetic correlations between adenomyosis and the same traits are shown on the y axis. The traits are colored by category.

Twenty-one traits were also genetically correlated with adenomyosis, while no significant correlation was observed for IBS, spontaneous abortion, and chronic fatigue (**Fig. 5** and **Supplemental Table 24**). The significant correlation between endometrial cancer and adenomyosis (rg=0.36, p=2.9×10^-3^) was absent with endometriosis (p=0.24). Considering different endometriosis definitions (i.e., combined, clinical, and self-reported), no phenotype showed a FDR-significant genetic correlation with preeclampsia or breast cancer. When comparing endometriosis clinical and self-reported definition (**Supplemental Table 24**), the latter had a stronger genetic correlation with anxiety (p_difference_=2.5×10^-5^) and MDD (p_difference_=3×10^-5^) than the former. Conversely, compared to self-reported definition, endometriosis clinical definition showed stronger genetic correlation with excessive, frequent, and irregular menstruation (p_difference_=2.1×10^-14^), female infertility (p_difference_=1.4×10^-3^), and endometriosis diagnosis and infertility diagnosis occurring together (p_difference_=2.5×10^-7^).

MixeR ^13^ was used to investigate further the pleiotropy between endometriosis (combined, clinical, and self-reported definition) and adenomyosis with genetically correlated traits (**Supplemental Table 25**). For instance, while the genetic correlation between endometriosis combined definition and MDD was 0.39, the fraction of shared variants was 95% with 98% of them with concordant effects. Evidence of strong pleiotropy was also observed between endometriosis clinical definition and female infertility (93% of shared variants with 92% of concordant effects within them). Interestingly, female-infertility pleiotropy was lower for endometriosis combined and self-reported definitions (62% and 37% of shared variants, respectively). Among other top results, endometriosis showed 91% of shared variants with “Excessive, frequent and irregular menstruation”. However, this estimate was lower when considering self-reported endometriosis (70% of shared variants). Mixer Bivariate models related to “female genital prolapse” showed good fits for the three endometriosis definitions and adenomyosis. Similar to the other phenotypes described above, “female male genital prolapse” shared the highest proportion of shared variants with endometriosis clinical definition (86%) compared to combined and self-reported ones (60% and 33%, respectively). Interestingly, with respect to this phenotype, adenomyosis shared the same proportion of shared variants observed for endometriosis clinical definition (86%). Different from the phenotypes described above, body mass index (BMI) showed a similar proportion of shared variants across endometriosis combined, clinical, and self-reported definitions (40%, 41%, and 41%, respectively). Nevertheless, within these shared variants, BMI showed 90% concordance with self-reported endometriosis, while only 79% and 75% were observed for the combined and clinical definitions, respectively.

Causal inference analyses using Mendelian randomization (MR) and latent causal variable (LCV) modeling revealed relationships linking endometriosis to several gynecological and systemic traits (**Fig. 6, Supplemental Tables 26** and **27**). Significant MR and LCV results converged on the effect of endometriosis on “excessive, frequent, and irregular menstruation” (MR β=1.3, p=2.6×10^−18^; LCV GCP=0.65, p=0.01). In the reverse MR analysis, a potential effect of this trait on endometriosis was shown, but the effect size was smaller (trait → endometriosis: β=0.15, p=1.2×10^−6^). Although these did not pass MR sensitivity analyses (see Methods), we observed other statistically significant evidence converging between MR and LCV analyses. These included the effect of endometriosis on “pain and other conditions associated with female genital organs and menstrual cycle” (endometriosis → trait: β=1.28, p=1.4×10^−14^; GCP=0.84, p=9.4×10^−6^) and “inflammatory disease of the uterus, excluding cervix” (MR β=0.36, p=2.4×10^−3^; LCV GCP=0.71, p=4×10^−3^) as well as the effect of leiomyoma of uterus on endometriosis (β=0.03, p=1.8×10^−3^; LCV GCP=0.42, p=1.8×10^−4^). LCV analysis also highlighted a possible effect of chronic fatigue on endometriosis (LCV GCP=0.74, p=1.8×10^−5^), but the lack of genetic instruments did not permit us to confirm this effect using MR (**Supplemental Table 26**).

**Figure 6.**
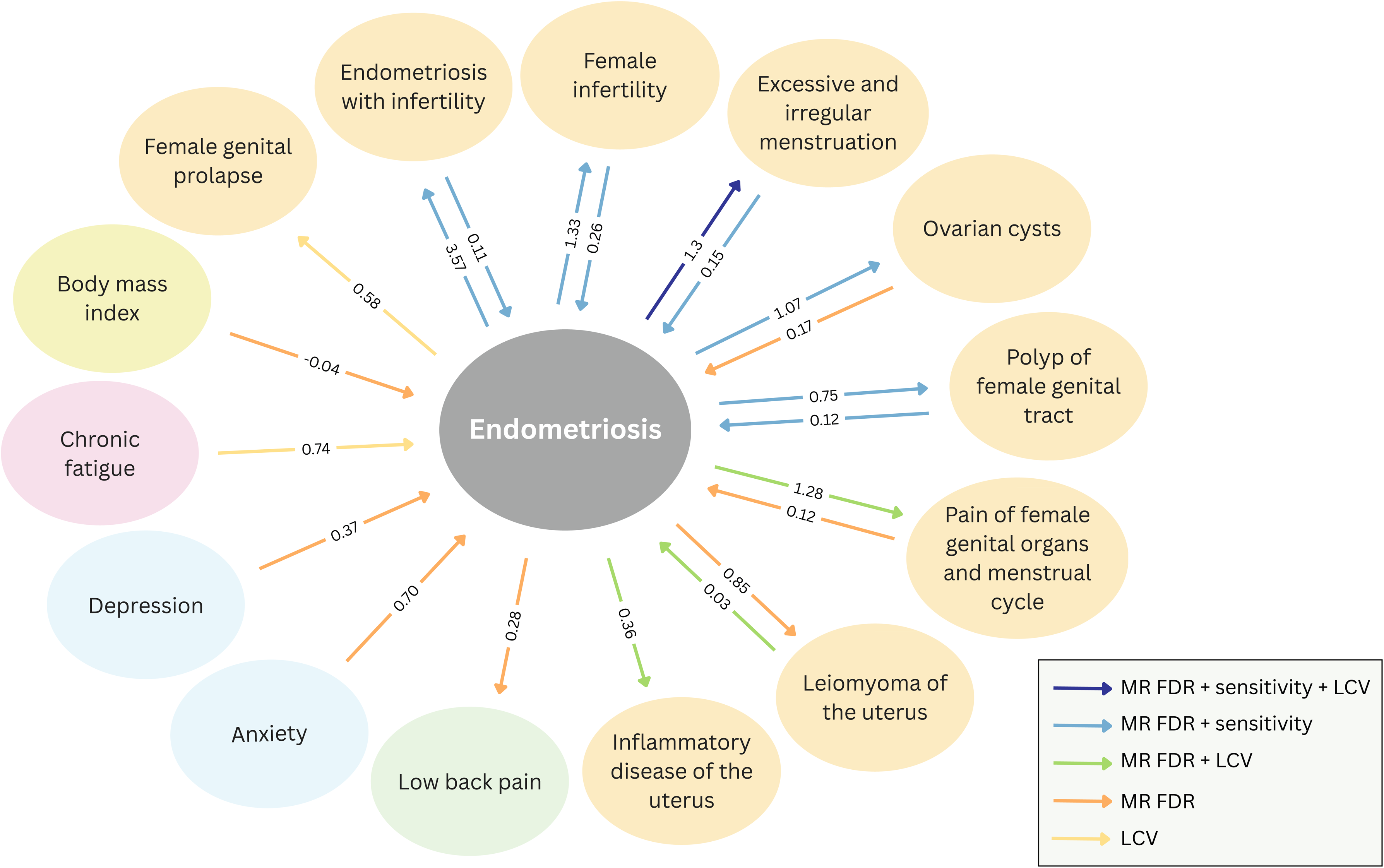
Genetically inferred causal relationships between endometriosis and gynecological/reproductive, pain-related, psychiatric, metabolic, and systemic traits. Traits are colored by category (gynecological/reproductive in orange, pain-related in green, psychiatric in blue, systemic in pink, and metabolic in yellow), and MR/LCV effect sizes are shown. The strength of the results is indicated with arrows, distinguishing results that survived FDR correction and sensitivity analyses in MR, results that survived FDR correction in MR and were supported by LCV, results significant after FDR correction in MR only, and results significant after FDR correction in LCV only.

## DISCUSSION

This study represents the largest multi-ancestry endometriosis GWAS to date, including approximately 1.4 million women, of whom 105,869 are cases, nearly doubling the number of cases included in previous GWAS ^4,7^. The cross-ancestry analysis including six population groups (i.e., AMR, AFR, EAS, EUR, MID, and SAS) increased the number of genome-wide significant loci for endometriosis from 63 in the EUR-only analysis to 80, of which 37 are novel. Additionally, we report the first five genome-wide significant loci for adenomyosis, two of which are shared with endometriosis and were not identified by previous GWAS ^4,7^.

Three of the five loci were not significant for endometriosis when adenomyosis cases were excluded, supporting their specificity. Genetic correlation analyses showed that endometriosis and endometriosis without adenomyosis had lower correlations with adenomyosis (rg = 0.79 and 0.81, respectively) than between different endometriosis definitions, supporting the differences between these conditions. Although adenomyosis has historically been referred to as ’endometriosis of the uterus’ , this terminology has been revised in ICD-11 ^14,15^. Still, the ongoing debate about whether they represent distinct diseases or phenotypic variations of a single disease continues ^16^. Furthermore, although we performed the first GWAS of adenomyosis excluding endometriosis cases, the reported prevalence of endometriosis in individuals with adenomyosis is 80.6%, and adenomyosis in individuals with endometriosis is 91.1%, suggesting a high degree of comorbidity and possible misclassification despite our efforts to separate these phenotypes ^17^.

Five loci were identified as genome-wide significant only in clinically confirmed endometriosis, despite the smaller sample size of this group. The previously found *ZHX3* was identified only for clinical endometriosis in our study, suggesting that the data source for endometriosis phenotyping can drive the specificity of gene discovery. Overall, our findings indicate that most GWAS signals are primarily driven by clinical endometriosis, as the majority of hits and fine-mapped variants showed the strongest evidence in this category.

This likely reflects more severe and symptomatic forms of the disease ^18^, as women correctly self-report endometriosis about 84% of the time ^18^. Our genetic correlation analysis showed a high rg=0.85 between clinical and self-reported endometriosis, consistent with phenotypic data, supporting the use of self-reported information in large-scale endometriosis studies ^18^.

For the first time, we also demonstrate that European-derived PRS is statistically associated with endometriosis across population groups. While its predictive power is currently insufficient for diagnostic use, they highlight the need to develop ancestry-informed risk models and to integrate genetic risk with clinical features.

Our integrative multi-omic analyses provide compelling evidence for shared genetic mechanisms influencing endometriosis risk through tissue-specific regulation of gene expression, protein levels, and DNA methylation. The functionally convergent results (colocalization, TWAS, and mQTL-based SMR) were observed for *VEZT*, *CDC42*, and *WNT4*. These loci were identified by previous endometriosis GWASs ^4,7^, but there was limited information regarding the functional mechanisms underlying their associations. Our study provided additional evidence for *VEZT* (rs9739834) through colocalization in the sigmoid colon. We also identified colocalization signals in novel loci, such as *HYAL3* rs13100173 across multiple reproductive and intestinal tissues, and *ATF1* rs61926299 in the ovary, colon, and breast mammary tissues. *HYAL3* has been identified as a shared gene between endometriosis and osteoarthritis, supporting the involvement of connective tissue remodeling pathways ^19^. *ATF1* may contribute to endometrial cyst formation through its interaction with steroid receptor RNA activator protein (SRAP) ^20^. In EAS, rs2388493 colocalized with *BAALC* transcriptomic regulation in sigmoid colon. To our knowledge, this is the first functional characterization of an endometriosis locus in non-EUR populations. Although *BAALC* is primarily studied in leukemia, it plays a role in cell proliferation, differentiation, and survival ^21^, which may reflect an EAS-specific mechanism in endometriosis pathophysiology.

Our TWAS results complement the colocalization findings by identifying 41 genes whose genetically predicted expression was significantly associated with endometriosis risk across multiple relevant tissues, compared to the 11 genes identified in the previous study that focused on uterine and blood tissues ^7^. This multi-tissue approach captures the complex biology of endometriosis, involving reproductive tissues such as uterus, ovary, and vagina, but also breast mammary tissue and colon, reflecting the systemic nature of the disease and its symptoms ^1^. Our PWAS identified nine significant proteins, compared to one, RSPO3 in a previous study ^7^, which association we replicated. *DTD1* showed a positive association with endometriosis in TWAS, but a negative association in PWAS. This discrepancy may reflect post-transcriptional regulation and tissue-specific differences, where higher mRNA levels might support ectopic cell survival, while higher actual protein levels confer protective effects by maintaining translational fidelity ^22^. We replicated seven previously reported TWAS associations ^7^, but observed variation in the tissues showing the strongest effects, which may also reflect the systemic nature of endometriosis ^1^. The novel genes and proteins we identified are involved in cell structure ^23^ and glycosylation ^24^ (e.g., *WASHC3*, *GMPPB*, *ABO*), which may promote ectopic cell survival and invasiveness, as previously hypothesized^25^. Immune-related genes (*SKAP1*, *HLA-DRA*, PRSS57) suggest impaired immune surveillance and inflammation ^26^. Several genes (*HOXA3/4*, *MST1*, *E2F6*, *CRISPLD1*) point to disrupted developmental signaling ^27^, autophagy ^28^, and cell cycle control ^29^, potentially contributing to lesion formation. SHBG and SAT2 may promote an estrogen-dominant ^30^ and proliferative ^31^ environment, respectively. Lastly, tumor suppressor and extracellular matrix-related genes and proteins (*RBM6*, FN1, CD109, CYB5D2, MANBA) showed protective effects when upregulated, underscoring the importance of structural integrity ^32^ and growth control ^33^. Our SMR meta-analyses further validated key loci in blood and endometrial tissue, replicating known genes (*VEZT*, *ESR1*, *WNT4*), and identified over 300 and 50 novel CpG associations in blood and endometrium, respectively, including *CBX3*, *SNX10*, *HNRNPA2B1*, and several loci in the MHC region. Some SMR signals appeared to be specific to different endometriosis definitions (clinical vs. self-reported), supporting molecular heterogeneity within the disease. These findings highlight novel immunological and inflammatory ^34,35^, and signaling ^36^ mechanisms. The pathway and gene-set enrichment analyses, which identified 1,727 regions, further confirmed the importance of immune regulation ^26^, tissue remodeling ^37^, and cell differentiation ^38^ in endometriosis. The overlap between endometriosis and adenomyosis suggests shared etiological components, also consistent with their clinical co-occurrence. However, the greater enrichment in epithelial cell proliferation and differentiation in endometriosis may reflect to tissue-specific mechanisms and differences in disease progression ^16^. Overall, our multi-tissue, multi-definition approach validated known associations and uncovered numerous novel genetic, proteomic, and epigenetic loci, offering a more comprehensive view of the genes, proteins, and regulatory mechanisms involved in endometriosis pathophysiology.

Recent studies have highlighted new therapeutic targets and repurposing opportunities for endometriosis, including immune-related genes (e.g., *GBP2*, *HCK*) ^39^. Our genetically informed analyses identified five promising repurposed drugs, including three breast cancer agents (neratinib, toremifene, abemaciclib). Toremifene, a selective estrogen receptor modulator, has been studied for its effects on the uterus and vagina, demonstrating similar estrogenic activity in both tissues and raising the possibility of off-target impacts on endometrial tissue ^40^. Similarly, neratinib, an EGFR/HER2 inhibitor, has been shown to be a potential treatment option for certain patients with endometrial cancer ^41^. The two progestins (17-hydroxyprogesterone caproate and norgestimate) are not currently used for endometriosis treatment but may help manage symptoms directly or indirectly related to the condition, such as heavy menstrual bleeding or preterm birth. Additional enrichment pointed to KIT inhibitors, GABA receptor antagonists, antipsychotics, and hypnotics. These insights emphasize shared neuroimmune, hormonal, and pain-related mechanisms, rather than pharmacological interventions directly targeting endometrial-like tissue.

We also observed a strong genetic overlap of endometriosis with gynecological/reproductive, psychiatric, pain-related, metabolic, gastrointestinal, cancer, and systemic traits. MixeR analyses further confirmed these relationships, with up to 95% of endometriosis-influencing variants shared with MDD, and similarly high overlap with pain and menstrual-related traits, supporting shared neuroendocrine and inflammatory mechanisms ^42,43^. In contrast, few variants were shared with cervical cancer. Adenomyosis showed a unique correlation with endometrial cancer, unlike endometriosis. Self-reported cases exhibited stronger pleiotropy with traits like BMI and ectopic pregnancy, while clinical and self-reported definitions showed similar overlap with infertility. These patterns underscore the importance of precise phenotyping to capture heterogeneity in endometriosis-related genetic architecture. The strongest causal relationship supported by both MR and LCV analyses was between endometriosis and excessive, frequent, and irregular menstruation. This provides robust genetic support for the long-standing hypothesis that menstrual dysregulation plays a key role in endometriosis pathogenesis ^44^. The genetic architecture of endometriosis may also predispose individuals to abnormal uterine bleeding patterns, potentially through shared mechanisms such as inflammatory dysregulation, impaired endometrial apoptosis, or altered matrix remodeling, findings also supported by our multi-omic analyses. This is further supported by a causal effect of uterine leiomyoma on endometriosis and bidirectional relationships with genital tract polyps, both conditions commonly comorbid with abnormal uterine bleeding (30% ^45^ and 50% ^46^, respectively). We also observed bidirectional associations between endometriosis and infertility, consistent with previous genetic studies ^4^ and the 30–50% comorbidity reported clinically ^47^. We identified a causal effect of endometriosis on ovarian cysts for the first time, aligning with clinical data showing their presence in 17–44% of women with endometriosis ^48^.

Our results provide the first genetic evidence suggesting a causal effect of chronic fatigue on endometriosis, consistent with prior observational data showing that over 50% of women with endometriosis experience fatigue, which is strongly associated with insomnia, depression, and pain ^49^. This supports the idea that fatigue is not merely a comorbidity, but may share neuroimmune or neuroendocrine pathways with endometriosis, further reflecting the systemic nature of the disease ^49^. Finally, a negative causal effect of BMI on endometriosis was observed, replicating previous MR studies ^50^ and epidemiological findings that suggest lower BMI may predispose individuals to endometriosis ^51^. Collectively, these findings underscore the complex, partly bidirectional relationships between endometriosis and gynecological and systemic conditions, and highlight the utility of genetic causal inference in uncovering previously unconfirmed links. Complementing these findings, our symptom-based PRS interaction analyses suggest that genetic risk may attenuate in individuals with high systemic symptom burden and abdominal pain, anxiety, migraine, and nausea. In line with our findings, a recent study reported that among women with endometriosis, those with a higher comorbidity burden had lower genetic predisposition ^52^. Hence, in the context of extensive symptomatology, non-genetic factors may play an important role in disease manifestation.

Our findings offer genetic and molecular support for several leading theories of endometriosis pathogenesis, including benign metastasis, dysregulated immune system, cell cycle and differentiation, impaired apoptosis, inflammation, hormonal imbalance, vascular dissemination, and epigenetic alterations ^53^. These results suggest that the complex pathophysiology of endometriosis may be due to the interplay of several biological mechanisms, rather than a single causal pathway, highlighting the need for integrative approaches to fully understand this complex disease.

## METHODS

### Study Populations

The GWAS meta-analysis included eight cohorts, UKB ^54^, AoU ^55^, MVP ^56^, BBJ ^57^, the International Endogene Consortium ^4^, FinnGen ^58^, EstBB ^59^, and 23andMe ^60^ (Supplemental Table 1). As previously collected, deidentified data were used in the present study, no ethical approval was needed ^61^. The study was additionally approved by the Norwegian Regional Committees for Medical and Health Research Ethics (REK 467496). The original cohorts were all approved by their institutional review boards and all participants provided written informed consent. In total, our cross-ancestry meta-analysis included 105,869 cases and 1,282,731 female controls (**Supplemental Table 1**).

UKB is a large population-based study that collected genetic, health, and lifestyle information from over 500,000 participants across the United Kingdom ^54^. Self-reported (field code 1402) and clinical endometriosis (ICD-10: N80) were included in the analyses. Nine GWASs were performed: ancestry-specific GWASs for AFR, AMR, CSA, EAS, MID, and EUR from medical records, EUR from self-reported phenotype information, EUR with only adenomyosis cases (“endometriosis of uterus”, ICD-10: N80.0), and EUR endometriosis cases without adenomyosis (ICD-10: N80.1-N80.9; **Supplemental Table 1**). The EUR GWAS of endometriosis combined definition was included in the International Endogene Consortium meta-analysis, hence it was not performed. The PLINK 2 ^62^ Firth regression (i.e., a penalized method that reduces bias when data are sparse or unbalanced) was used for each GWAS to estimate genetic associations with covariates including age and the first ten within-ancestry principal components (PCs). The following quality control criteria were applied: biallelic variants with minor allele frequency > 1%, Hardy–Weinberg equilibrium P < 10^−6^, variant call rate > 95%, and per-individual genotyping rate > 95%. Genetically inferred ancestry assignment was derived from the Pan-UKB Initiative ^63^.

The AoU Research Program is an ancestrally diverse US-based precision medicine initiative aiming to enroll over one million participants to reflect the diversity of the population, with rich genomic, clinical, and survey data ^55^. Ten GWASs were performed using the same approach as for UKB, with the addition of a meta-analysis within the EUR group. Genetic associations were estimated using PLINK 2 ^62^ logistic regression, including age and the first ten within-ancestry PCs as covariates. The same quality control criteria were applied as in the UKB analysis. AoU genetically inferred ancestry assignment was previously described ^55^.

MVP is a U.S. Department of Veterans Affairs cohort that includes genetic and health data from over 900,000 U.S. military veterans to study the genetic basis of diseases ^56^. The GWAS data used in this study were generated from 16,231 and 32,192 female individuals of AFR and EUR ancestry, respectively. The genetic association analysis was performed using a PLINK 2 ^62^ logistic regression model with covariates for age, sex, and the first ten within-ancestry PCs. Details of MVP GWAS have been previously described ^64^.

BBJ is a hospital-based biobank that collected DNA and clinical data from over 200,000 participants across Japan, focused on a wide range of common diseases ^57^. The GWAS was conducted by employing a generalized linear mixed model (GLMM) using SAIGE ^65^, age, and top 5 PCs were used as covariates and it was generated from 82,761 female individuals of EAS ancestry.

The International Endogene Consortium is a collaborative effort that aggregates data from multiple international studies to investigate the genetic factors of endometriosis ^4^. GWAS data were generated from 474,160 individuals, of which 3,294 were of EAS ancestry. This meta-analysis included 23 cohorts with variable endometriosis case definition including medical records, surgically-confirmed endometriosis, and self-reported information, and most studies applied logistic regression. The covariates used varied: several analyses included no adjustment, while others adjusted for age, sex, and a range of PCs, from using the first two only and up to 20. The meta-analysis was performed using the inverse variance weighting fixed-effects method implemented in METAL ^66^.

FinnGen is a public-private partnership combining genetic data from over 500,000 Finnish biobank participants with nationwide health registry data to advance understanding of disease genetics ^58^. The FinnGen GWAS was performed using REGENIE ^67^, including covariates for age, sex, the top ten within-ancestry PCs, and genotyping batch. The GWAS data for endometriosis (ICD10: N80) and adenomyosis (ICD10: N80.0) were derived from Release 12, including 150,350 and 135,833 EUR individuals of EUR ancestry, respectively.

EstBB is a population-based cohort comprising over 200,000 individuals from Estonia, with comprehensive genetic and health data linked to national health records ^59^. The EstBB GWAS was performed using REGENIE ^67^, including covariates for year of birth and the top ten within-ancestry PCs. Endometriosis GWAS data (ICD10: N80) were generated from 126,807 individuals of EUR ancestry.

23andMe is a direct-to-consumer genetics and research company with a cohort of research-consented customers, totaling millions of individuals, contributing genetic and self-reported health data to research ^60^. The 23andMe endometriosis GWAS was performed using logistic regression models with current age, the top five PCs, and the genotyping platform as covariates ^60^.

Given the different endometriosis phenotype definitions, we compared the genetic architecture between different GWASs using LD score regression ^68^. LD scores were calculated using HapMap 3 variants ^69^ and the 1000 Genomes Project EUR reference population ^70^. These analyses were only performed in EUR due to the low sample size of other ancestry-specific GWASs.

### Ancestry-Specific and Cross-Ancestry Meta-Analyses

Before meta-analyzing the different datasets, the following quality control steps were applied: minor allele frequency > 1%, imputation INFO score > 0.8 when available, and exclusion of non-rs IDs and non-biallelic variants. For the cross-ancestry meta-analysis, the fixed-effects inverse variance weighting method implemented in METAL ^66^ was used, based on sample sizes and P-values to account for possible sample overlap, resulting in 39,620,082 SNPs.

After meta-analysis, we excluded variants that were not present in more than 50% of the total effective sample size (n = 195,595), resulting in 7,865,243 remaining SNPs. Additional meta-analysis were conducted for AFR (2,481 cases and 54,671 controls), AMR (1,752 cases and 37,871 controls), EAS (2,044 cases and 87,723 controls), EUR (99,407 cases and 1,093,534 controls), EUR clinical (36,695 cases and 567,775 controls), EUR self-reported (50,867 cases and 528,391 controls), adenomyosis (8,753 cases and 415,718 controls), and endometriosis without adenomyosis (6,627 cases and 283,316 controls). LD score regression metrics, including SNP-*h*^2^, genomic inflation (lambda), mean chi-squared statistic, LDSC intercept, and confounding ratio, were calculated for each meta-analysis.

We performed a joint analysis using GCTA-COJO ^71,72^ to identify secondary association signals in the loci reaching genome-wide significance with respect to cross-ancestry, EUR, and EAS GWAS of endometriosis combined definition and of EUR clinical definition. A stepwise model selection procedure was applied to identify independently associated SNPs. We used UKB ancestry-specific genome-wide data as the linkage disequilibrium (LD) reference panel. The default parameters were considered: 10 Mb for LD distance; 0.9 for multiple regression *R*^2^ cutoff for collinearity; 0.2 for difference in allele frequency between the GWAS and LD reference. SNPs with joint-analysis p-value (P_J_)<5×10^−8^ were considered genome-wide significant. Among the SNPs identified by COJO, we further excluded those with LD r^2^≥0.1 with any lead SNPs identified in the primary GWAS.

### Functional Characterization

#### Fine-mapping and colocalization

To identify causal variants underlying the GWS associations, we conducted fine-mapping using PAINTOR (version 3), a probabilistic framework that incorporates functional annotations to model their effects on the magnitude of SNP associations and prioritize likely causal variants ^73^. We analyzed LD-independent endometriosis-associated loci in EUR, EUR-clinical, EAS, and cross-ancestry datasets, using a ±1 Mbp window around the lead SNP at each locus. To select relevant functional annotations, we first estimated the enrichment of endometriosis heritability in EUR, EUR-clinical, EAS, and cross-ancestry GWAS using partitioned heritability analysis via LDSC and the baseline annotation set ^74^. Among the Bonferroni-significant annotations (N_tests_=97, p<5×10⁻⁴), we retained those for which endometriosis-specific functional data were available in PAINTOR. To address multicollinearity among annotations, we clustered the annotation matrices separately for each ancestry-specific summary statistic. From each cluster, we selected a representative annotation—prioritizing distinct functional categories and maximizing dissimilarity from other clusters where possible. Clustering and variable selection were performed in R using the klaR package. Fine-mapping was conducted without pre-specifying the number of causal variants, using PAINTOR’s Gibbs sampling algorithm to sample from the posterior distribution directly.

We conducted colocalization analysis to identify shared causal SNPs between endometriosis GWAS and transcriptomic regulation data from six endometriosis-relevant tissues available from GTEx v8 (i.e., uterus, ovary, vagina, breast mammary, and sigmoid and transverse colon) ^75^. Specifically, considering default parameters, we used HyPrColoc deterministic Bayesian algorithm to estimate the PPFC ^76^. The colocalization analysis was conducted with respect to cross-ancestry, EUR, and EAS GWAS of endometriosis combined definition and EUR clinical definition. Following developers’ recommendation ^76^, we used PPFC>0.7 as the threshold for identifying colocalization signals. In addition, we prioritized candidate causal SNPs with the threshold of proportion of the PPFC explained by the candidate causal variant (P_snp_>0.5). Estimates of regional association probability (P_R_) were also calculated.

#### Transcriptome- and proteome-wide analyses

Tissue-specific and cross-tissue TWAS of EUR endometriosis combined definition were performed using S-PrediXcan and S-MultiXcan approaches (available at https://github.com/hakyimlab/MetaXcan) ^77,78^. Leveraging GTEx v8 data ^75^, tissue-specific predictive models of gene expression and their corresponding LD covariance matrices were previously calculated (available at https://predictdb.org/categories/downloads/). We restricted the TWAS to seven tissues relevant to the pathogenesis of endometriosis and its symptoms: uterus, vagina, ovary, breast mammary tissue, whole blood, colon sigmoid, and colon transverse. Bonferroni multiple testing correction was applied to account for the number of genes tested (N=13,457; p<3.72×10^-6^).

We conducted a PWAS to identify genes whose cis-regulated plasma protein abundance is associated with endometriosis risk. The analysis was performed using FUSION ^79^, which integrates genome-wide association statistics with pre-trained protein imputation models. We specifically used the elastic-net (enet) models, as recommended by the method developers, due to their higher prediction accuracy compared to other approaches. In the present study, the PWAS was conducted using the endometriosis GWAS meta-analysis including individuals of European ancestry. We used protein quantitative trait locus (pQTL) weights derived from the plasma proteome study conducted by Zhang et al. ^80^, which includes imputation models for 4,657 plasma proteins trained in 7,213 individuals of European ancestry ^70^. From these, imputation models were available for 1,318 plasma proteins with significant SNP-*h^2^* (p<0.01) and were included in our PWAS analysis. The European subset of the 1000 Genomes Project ^70^ was used as the reference panel. We applied a false discovery correction (FDR q<0.05) accounting for the number of proteins tested.

#### mQTL-based SMR

We used the SMR approach ^81^ to assess the effect of methylation changes on endometriosis and adenomyosis. Specifically, we integrated information regarding mQTL in blood (up to 256,524 CpG sites) and endometrial tissue (3,034 CpG sites) with genome-wide association statistics generated by the present study. The analysis was performed using ancestry-stratified information using matching reference populations available from the 1000 Genomes Project Phase 3. With respect to EUR, blood mQTL datasets included the GoDMC catalog ^82^, the ARIES project ^83^, Hatton et al ^84^, McRae et al ^85^, Hannon et al 2018 ^86^, and Hannon et al 2016 ^87^. In EUR, we also investigated an endometrium-specific mQTL dataset ^88^. A blood mQTL dataset was also available for EAS^86^. No other population-specific mQTL dataset was available for the other ancestry groups investigated in the present study. SMR analyses were limited to cis-mQTLs (±2 Mb of each CpG site), with thresholds applied for SMR testing (p<5×10⁻⁸), LD pruning (r² between 0.05 and 0.9), and the HEIDI test (p>0.05 using up to 20 top cis-SNPs). To enhance statistical power, in EUR blood-specific SMR results were meta-analyzed using sample size-weighted METAL ^66^, applying genomic control correction and sample overlap adjustment enabled via Z-statistic–based estimation. HEIDI test p > 0.05 in each of the datasets meta-analyzed was considered to exclude biases due to heterogeneity. False discovery rate was applied to account for the number of CpG sites tested (FDR q<0.05).

### Drug Repurposing

We used a drug repositioning approach, DRUGSETS (https://github.com/nybell/drugsets). Data were extracted from the Drug Gene Interaction Database and the Clue Repurposing Hub. A total of 1,201 drug-gene sets were defined for medications based on genes whose protein products are either direct molecular targets or known interactors with the respective drug. We performed competitive gene set analysis using MAGMA v.1.10 ^89^, conditioning on a background gene set consisting of 2,281 drug target genes, to test for significant associations between drug–gene sets and endometriosis. Bonferroni correction was applied for multiple testing based on the number of drug-gene sets tested (n=735, p<6.80×10^−5^).

Drug-gene sets were subsequently classified in three ways: by Anatomical Therapeutic Classification (ATC) III code, by clinical indication, and by the mechanism of action (MOA). A multiple linear regression approach was used to test whether membership in each drug group predicted stronger gene set associations, with group membership as the predictor and the MAGMA drug gene-set t-statistic as the outcome.

### Gene Ontology Enrichment

To characterize endometriosis pathogenesis, we performed a gene-set enrichment analysis using the GSA-MiXeR framework (available at https://github.com/precimed/gsa-mixer). This allows to quantify heritability enrichments with respect to predefined gene sets while accounting for LD structure ^90^. The primary GSA-MiXeR analysis was performed with respect to EUR GWAS meta-analysis of endometriosis combined phenotypes. We also investigated EUR adenomyosis to assess differences between these comorbid conditions.

Additionally, we also assess EUR endometriosis clinical definition and EUR endometriosis self-reported definition. EUR populations available from 1,000 Genomes Project were used as LD reference panel ^70^. Gene sets were defined based on Gene Ontology (GO) annotations, including biological processes (BP), molecular functions (MF), and cellular components (CC). Bonferroni correction was applied to account for the number of GOs tested (N=79,223; p<6.31×10⁻⁷). To reduce redundancy in GO term enrichments and identify representative functional themes, we employed REVIGO ^12^, using a semantic similarity cutoff of 0.7 and UniProt as the reference database.

### Polygenic Risk Scoring

To investigate how endometriosis polygenic risk translates across ancestry groups, we derived PRS from the EUR endometriosis combined definition with and without the AoU cohort, and tested them in all populations in AoU, that is AFR, AMR, EAS, EUR, MID, and SAS. For the EUR analysis, we used summary statistics that excluded the AoU cohort.

Posterior variant-level effect sizes for the EUR endometriosis combined definition were estimated using PRS-CS ^91^, with the 1000 Genomes Project European population as the LD reference panel ^70^. Endometriosis PRS were then computed using PLINK2 ^62^, then the PRS estimates were normalized. Additionally, we evaluated the association of PRS quartiles with disease risk using logistic regression models, treating the first quartile as the reference, within each AoU ancestry group.

PRS were calculated in the same manner for UKB endometriosis cases and controls of EUR ancestry, using effect sizes derived from the EUR endometriosis combined definition, excluding the International Endogene Consortium data which includes UKB. Using data from EUR individuals in both AoU and UKB, we conducted a PRS interaction analysis using a logistic regression model to assess whether the association between the endometriosis PRS and endometriosis risk differs depending on the presence or absence of endometriosis systemic symptoms and comorbidities (i.e., self-reported and medical diagnoses of abdominal pain, anxiety, bloating, chronic pelvic pain, constipation, contraceptive use, depression, diarrhea, dysmenorrhea, dyspareunia, dysuria, eating disorders, fatigue, fibromyalgia, irritable bowel syndrome, infertility, length of menstrual cycle, low back pain, menorrhagia, migraine, nausea, pain severity, and the sum of all traits, comorbidity burden; **Supplemental Table 22**). Age at recruitment and the top-10 within-ancestry PCs were added as covariates for all analyses.

### Pleiotropy Analysis

#### Genetic correlation

Global rg was calculated between EUR endometriosis (combined, self-reported, and clinical endometriosis, and adenomyosis) and 35 traits related to female health in seven categories (gynecological/reproductive, psychiatric, pain-related, metabolic, gastrointestinal, cancer, and systemic; **Supplemental Table 23**) were conducted using the Linkage Disequilibrium Score Regression (LDSC) method v1.0.1. ^68^. Effective sample sizes ranged from 5,632 to 1,816,976. To ensure reliable estimates, the genetic correlation analysis was limited to traits with a SNP-*h^2^* p<0.05 (**Supplemental Table 24**). An FDR multiple testing correction (FDR q<0.05) was applied to define statistically significant genetic correlations. To evaluate heterogeneity across definitions, meta-analyzed endometriosis was compared to adenomyosis, and self-reported endometriosis was compared to endometriosis by clinical definitions using a Z-test. We also used MiXeR mixture models ^13^ to estimate the number of shared variants between endometriosis and traits with FDR-significant global genetic correlations.

#### Mendelian randomization

We conducted a bidirectional two-sample MR analysis implemented in the TwoSampleMR R package ^92^ to investigate the potential causal relationship between endometriosis and genetically correlated traits (q<0.05). The primary method used was the multiplicative random-effects inverse variance weighted (IVW) approach ^93^. To avoid sample overlap, GWAS summary statistics excluding the shared sample were used for each trait. Only independent SNPs (r²<0.001 in a 10,000 kb window) with genome-wide significance (P<5×10^-8^) in the exposure were included. SNPs that reached significance in both the exposure and the outcome were excluded. Potential causal relationships were removed from the analyses when only one independent significant SNP remained as a valid instrument. P-values were adjusted for multiple comparisons using the FDR correction.

For IVW results with q<0.05, we undertook sensitivity analyses to assess the robustness of the findings. The weighted median and weighed mode methods were used since they are less sensitive to outliers and remain reliable even when only a portion of the genetic variants are valid instruments ^94,95^. The MR-PRESSO global test was used to remove horizontal pleiotropic outliers and, if detected, the IVW method was rerun using the remaining instruments ^96^. To minimize the risk of reverse causation, Steiger filtering was used to exclude instruments that explained more variance in the outcome than in the exposure, and the IVW method was repeated on the remaining set of instruments ^97^. Additionally, if there was a sign of pleiotropy (p-value from MR-Egger intercept<0.05), the I^2^ statistic was calculated to quantify the strength of the violation of the NO Measurement Error (NOME) assumption ^98^. If the NOME assumption was not violated (I^2^>0.9), the MR-Egger was applied, since it allows all genetic variants to have pleiotropic effects ^99^. When 0.6<I^2^< 0.9, we applied the simulation extrapolation (SIMEX) method to generate an adjusted causal effect estimate ^98^. When I^2^ < 0.6, MR-Egger results were not taken into consideration. Evidence of a causal relationship was considered if the same direction of effect as the IVW beta estimate and p<0.05 was found for the (i) weighted median method, (ii) weighted mode method, (iii) IVW after MR PRESSO, (iv) IVW after Steiger filtering, and (v) in case of pleiotropy (MR-Egger intercept p<0.05), MR-Egger or SIMEX if I^2^>0.9 or 0.6<I^2^< 0.9, respectively.

#### Latent causal variable analysis

To triangulate causal evidence, we performed a LCV analysis using LD scores derived from European populations in phase 3 of the 1000 Genomes Project. LCV provided estimates of the Genetic Causal Proportion (GCP), where the sign of the GCP indicates the direction of causality: positive values suggest that phenotype 1 influences phenotype 2, and negative values indicate the reverse. In the present study, we presented only positive GCP values, specifying the direction of the effect they correspond to. The direction of the LCV effect is reflected by the ρ statistic, with ρ>0 indicating a positive effect and ρ < 0 indicating a negative effect. FDR q<0.05 was considered to define significant results after multiple testing correction.

## Data Availability

The genome-wide association statistics generated by the current study will be made available in Zenodo at the time of publication.

## ACKNOWLEDGMENTS

The authors acknowledge support from the National Institutes of Health (RF1 MH132337 to R.P.), the American Foundation for Suicide Prevention (PDF-0-065-23 to J.H.), the MQ Foundation (UFA21\100014 to B.C.M.), ’Fundació La Marató de TV3’ (202218-31 to B.C.), the Spanish ‘Ministerio de Ciencia, Innovación y Universidades’ (projects PID2021-1277760B-I100 and PID2024-158634OB-I00 funded by MICIU/AEI/10.13039/501100011033/ and FEDER-EU to B.C.; PID2022-139740OA-I00 funded by MICIU/AEI/10.13039/501100011033/ and FEDER-EU; RYC2021-033573-I funded by MCIN/AEI/10.13039/501100011033 and by the European Union “NextGenerationEU”/PRTR to M.M.; RYC2024-050099-I and JDC2024-055161-I funded by MICIU/AEI/10.13039/501100011033 and by FSE+ to D.K. and S.A., respectively; and the Endo-Map project PID2021-12728OB-I00 funded by MICIU/AEI/10.13039/501100011033 and by FEDER-EU to S.A.)), ICREA Academia 2021 (to B.C.), AGAUR (2021SGR-01093 to B.C. and M.M.), and the University of Bergen (International Training Grant to S.L.).We also acknowledge the contribution of the participants and the investigators involved in the UKB, the FinnGen Project, the MVP, the AoU Research Program, the EstBB, BBJ, and all included studies in the International Endogene Consortium GWAS. We would like to thank the research participants and employees of 23andMe, Inc. for making this work possible. The research using UKB resources has been conducted under Application Number 58146. The AoU Research Program is supported by the National Institutes of Health, Office of the Director: Regional Medical Centers: 1 OT2 OD026549; 1 OT2 OD026554; 1 OT2 OD026557; 1 OT2 OD026556; 1 OT2 OD026550; 1 OT2 OD 026552; 1 OT2 OD026553; 1 OT2 OD026548; 1 OT2 OD026551; 1 OT2 OD026555; IAA: AOD 16037; Federally Qualified Health Centers: HHSN 263201600085U; Data and Research Center: 5 U2C OD023196; Biobank: 1 U24 OD023121; The Participant Center: U24 OD023176; Participant Technology Systems Center: 1 U24 OD023163; Communications and Engagement: 3 OT2 OD023205; 3 OT2 OD023206; and Community Partners: 1 OT2 OD025277; 3 OT2 OD025315; 1 OT2 OD025337; 1 OT2 OD025276.

## REFERENCES

1. Taylor, H. S., Kotlyar, A. M. & Flores, V. A. Endometriosis is a chronic systemic disease: clinical challenges and novel innovations. The Lancet 397, 839–852 (2021).

2. Missmer, S. A. et al. Impact of Endometriosis on Life-Course Potential: A Narrative Review. IJGM **Volume** 14, 9–25 (2021).

3. Saha, R. et al. Heritability of endometriosis. Fertility and Sterility 104, 947–952 (2015).

4. Rahmioglu, N. et al. The genetic basis of endometriosis and comorbidity with other pain and inflammatory conditions. Nat Genet 55, 423–436 (2023).

5. Koller, D. et al. Epidemiologic and Genetic Associations of Endometriosis With Depression, Anxiety, and Eating Disorders. JAMA Netw Open 6, e2251214 (2023).

6. Sapkota, Y. et al. Meta-analysis identifies five novel loci associated with endometriosis highlighting key genes involved in hormone metabolism. Nat Commun 8, 15539 (2017).

7. Guare, L. A., et al. Expanding the genetic landscape of endometriosis: Integrative -omics analyses uncover key pathways from a multi-ancestry study of over 900,000 women. Preprint at 10.1101/2024.11.26.24316723 (2024).

8. Moawad, G. et al. Adenomyosis: An Updated Review on Diagnosis and Classification. J Clin Med 12, 4828 (2023).

9. International Working Group of AAGL, ESGE, ESHRE and WES et al. Endometriosis classification, staging and reporting systems: a review on the road to a universally accepted endometriosis classification. Facts Views Vis Obgyn 13, 305–330 (2021).

10. Gerber, Z., Fisun, M., Aschard, H. & Djebali, S. PaintorPipe: a pipeline for genetic variant fine-mapping using functional annotations. Bioinformatics Advances 4, (2024).

11. Bell, N., Uffelmann, E., Van Walree, E., De Leeuw, C. & Posthuma, D. Using genome-wide association results to identify drug repurposing candidates. Preprint at 10.1101/2022.09.06.22279660 (2022).

12. Supek, F., Bošnjak, M., Škunca, N. & Šmuc, T. REVIGO summarizes and visualizes long lists of gene ontology terms. PLoS One 6, e21800 (2011).

13. Frei, O. et al. Bivariate causal mixture model quantifies polygenic overlap between complex traits beyond genetic correlation. Nat Commun 10, (2019).

14. Harrison, J. E., Weber, S., Jakob, R. & Chute, C. G. ICD-11: an international classification of diseases for the twenty-first century. BMC Med Inform Decis Mak 21, 206 (2021).

15. Whitaker, L. H. R. et al. Proposal for a new ICD-11 coding classification system for endometriosis. Eur J Obstet Gynecol Reprod Biol 241, 134–135 (2019).

16. Habiba, M., Guo, S.-W. & Benagiano, G. Are Adenomyosis and Endometriosis Phenotypes of the Same Disease Process? Biomolecules 14, 32 (2023).

17. Leyendecker, G. et al. Adenomyosis and endometriosis. Re-visiting their association and further insights into the mechanisms of auto-traumatisation. An MRI study. Arch Gynecol Obstet 291, 917–932 (2015).

18. Shafrir, A. L. et al. Validity of self-reported endometriosis: a comparison across four cohorts. Human Reproduction 36, 1268–1278 (2021).

19. Shigesi, N. et al. The phenotypic and genetic association between endometriosis and immunological diseases. Hum Reprod 40, 1195–1209 (2025).

20. Onuma, T. et al. Zinc deficiency is associated with the development of ovarian endometrial cysts. Am J Cancer Res 13, 1049–1066 (2023).

21. Tanner, S. M. et al. BAALC, the human member of a novel mammalian neuroectoderm gene lineage, is implicated in hematopoiesis and acute leukemia. Proc Natl Acad Sci U S A 98, 13901–13906 (2001).

22. Yadavalli, S. S. & Ibba, M. Quality control in aminoacyl-tRNA synthesis its role in translational fidelity. Adv Protein Chem Struct Biol 86, 1–43 (2012).

23. Arendt, W., Kleszczyński, K., Gagat, M. & Izdebska, M. Endometriosis and Cytoskeletal Remodeling: The Functional Role of Actin-Binding Proteins. Cells 14, 360 (2025).

24. Kovács, Z. et al. N-glycans from serum IgG and total serum glycoproteins specific for endometriosis. Sci Rep 13, (2023).

25. Klemmt, P. A. B. & Starzinski-Powitz, A. Molecular and Cellular Pathogenesis of Endometriosis. Curr Womens Health Rev 14, 106–116 (2018).

26. Abramiuk, M. et al. The Role of the Immune System in the Development of Endometriosis. Cells 11, 2028 (2022).

27. Cho, Y. J. et al. Dysfunctional signaling underlying endometriosis: current state of knowledge. Journal of Molecular Endocrinology 60, R97–R113 (2018).

28. Yang, H.-L. et al. Autophagy in endometriosis. Am J Transl Res 9, 4707–4725 (2017).

29. D’Amora, P. et al. Disrupted cell cycle control in cultured endometrial cells from patients with endometriosis harboring the progesterone receptor polymorphism PROGINS. Am J Pathol 175, 215–224 (2009).

30. Misao, R., Hori, M., Ichigo, S., Fujimoto, J. & Tamaya, T. Levels of sex hormone-binding globulin (SHBG) and corticosteroid-binding globulin (CBG) messenger ribonucleic acid (mRNAs) in ovarian endometriosis. Reprod Nutr Dev 35, 155–165 (1995).

31. Le Page, C. et al. A COEUR cohort study of SATB2 expression and its prognostic value in ovarian endometrioid carcinoma. J Pathol Clin Res 5, 177–188 (2019).

32. Cook, C. J., Wiggin, N. & Fogg, K. C. Characterizing the Extracellular Matrix Transcriptome of Endometriosis. Reprod Sci 31, 413–429 (2024).

33. Rafique, S. & Decherney, A. H. Medical Management of Endometriosis. Clin Obstet Gynecol 60, 485–496 (2017).

34. Xiang, Y. et al. CBX3 antagonizes IFNγ/STAT1/PD-L1 axis to modulate colon inflammation and CRC chemosensitivity. EMBO Mol Med 16, 1404–1426 (2024).

35. You, Y. et al. Sorting nexin 10 acting as a novel regulator of macrophage polarization mediates inflammatory response in experimental mouse colitis. Sci Rep 6, (2016).

36. Lu, Y. et al. CRNDE mediated hnRNPA2B1 stability facilitates nuclear export and translation of KRAS in colorectal cancer. Cell Death Dis 14, (2023).

37. Yang, W.-C. V. et al. Matrix remodeling and endometriosis. Reprod Med Biol 4, 93–99 (2005).

38. Peinado, F. M. et al. Cell cycle, apoptosis, cell differentiation, and lipid metabolism gene expression in endometriotic tissue and exposure to parabens and benzophenones. Science of The Total Environment 879, 163014 (2023).

39. Liu, B. H. M. et al. Utilizing AI for the Identification and Validation of Novel Therapeutic Targets and Repurposed Drugs for Endometriosis. Advanced Science 12, (2025).

40. Tomás, E., Kauppila, A., Blanco, G., Apaja-Sarkkinen, M. & Laatikainen, T. Comparison between the effects of tamoxifen and toremifene on the uterus in postmenopausal breast cancer patients. Gynecol Oncol 59, 261–266 (1995).

41. Schwab, C. L. et al. Neratinib shows efficacy in the treatment of HER2/neu amplified uterine serous carcinoma in vitro and in vivo. Gynecol Oncol 135, 142–148 (2014).

42. Sherwani, S. et al. The vicious cycle of chronic endometriosis and depression-an immunological and physiological perspective. Front Med (Lausanne) 11, 1425691 (2024).

43. Vargas, E., Aghajanova, L., Gemzell-Danielsson, K., Altmäe, S. & Esteban, F. J. Cross-disorder analysis of endometriosis and its comorbid diseases reveals shared genes and molecular pathways and proposes putative biomarkers of endometriosis. Reproductive BioMedicine Online 40, 305–318 (2020).

44. Kuan, K. K. W., Gibson, D. A., Whitaker, L. H. R. & Horne, A. W. Menstruation Dysregulation and Endometriosis Development. Front Reprod Health 3, 756704 (2021).

45. Uimari, O., Subramaniam, K. S., Vollenhoven, B. & Tapmeier, T. T. Uterine Fibroids (Leiomyomata) and Heavy Menstrual Bleeding. Front Reprod Health 4, 818243 (2022).

46. Nijkang, N. P., Anderson, L., Markham, R. & Manconi, F. Endometrial polyps: Pathogenesis, sequelae and treatment. SAGE Open Med 7, 2050312119848247 (2019).

47. Endometriosis and Infertility: The Comorbidities. in Endometriosis-related Infertility 9–17 (Springer International Publishing, Cham, 2024). doi:10.1007/978-3-031-50662-8_2.

48. Matalliotaki, C. et al. Co-existence of benign gynecological tumors with endometriosis in a group of 1,000 women. Oncol Lett 15, 1529–1532 (2018).

49. Ramin-Wright, A. et al. Fatigue – a symptom in endometriosis. Human Reproduction 33, 1459–1465 (2018).

50. Garitazelaia, A. et al. A Systematic Two-Sample Mendelian Randomization Analysis Identifies Shared Genetic Origin of Endometriosis and Associated Phenotypes. Life (Basel) 11, 24 (2021).

51. Liu, Y. & Zhang, W. Association between body mass index and endometriosis risk: a meta-analysis. Oncotarget 8, 46928–46936 (2017).

52. McGrath, I. M., Rukins, V., Laisk, T., Mortlock, S. & Montgomery, G. W. Interaction between genetic risk and comorbid conditions in endometriosis. Human Genetics and Genomics Advances 6, 100456 (2025).

53. Lamceva, J., Uljanovs, R. & Strumfa, I. The Main Theories on the Pathogenesis of Endometriosis. Int J Mol Sci 24, 4254 (2023).

54. Bycroft, C. et al. The UK Biobank resource with deep phenotyping and genomic data. Nature 562, 203–209 (2018).

55. The All of Us Research Program Genomics Investigators et al. Genomic data in the All of Us Research Program. Nature 627, 340–346 (2024).

56. Gaziano, J. M. et al. Million Veteran Program: A mega-biobank to study genetic influences on health and disease. Journal of Clinical Epidemiology 70, 214–223 (2016).

57. Ishigaki, K. et al. Large-scale genome-wide association study in a Japanese population identifies novel susceptibility loci across different diseases. Nat Genet 52, 669–679 (2020).

58. Kurki, M. I. et al. FinnGen provides genetic insights from a well-phenotyped isolated population. Nature 613, 508–518 (2023).

59. Milani, L. et al. The Estonian Biobank’s journey from biobanking to personalized medicine. Nat Commun 16, 3270 (2025).

60. Galarneau, G. et al. Genome-wide association studies on endometriosis and endometriosis-related infertility. Preprint at 10.1101/401448 (2018).

61. Bathe, O. F. & McGuire, A. L. The ethical use of existing samples for genome research. Genetics in Medicine 11, 712–715 (2009).

62. Chang, C. C. et al. Second-generation PLINK: rising to the challenge of larger and richer datasets. Gigascience 4, 7 (2015).

63. Karczewski, K. J. et al. Pan-UK Biobank GWAS improves discovery, analysis of genetic architecture, and resolution into ancestry-enriched effects. Preprint at 10.1101/2024.03.13.24303864 (2024).

64. Verma, A. et al. Diversity and scale: Genetic architecture of 2068 traits in the VA Million Veteran Program. Science 385, eadj1182 (2024).

65. Zhou, W. et al. Efficiently controlling for case-control imbalance and sample relatedness in large-scale genetic association studies. Nat Genet 50, 1335–1341 (2018).

66. Willer, C. J., Li, Y. & Abecasis, G. R. METAL: fast and efficient meta-analysis of genomewide association scans. Bioinformatics 26, 2190–2191 (2010).

67. Mbatchou, J. et al. Computationally efficient whole-genome regression for quantitative and binary traits. Nat Genet 53, 1097–1103 (2021).

68. Bulik-Sullivan, B. K. et al. LD Score regression distinguishes confounding from polygenicity in genome-wide association studies. Nat. Genet. 47, 291–295 (2015).

69. International HapMap 3 Consortium et al. Integrating common and rare genetic variation in diverse human populations. Nature 467, 52–58 (2010).

70. The 1000 Genomes Project Consortium et al. A global reference for human genetic variation. Nature 526, 68–74 (2015).

71. Yang, J. et al. Conditional and joint multiple-SNP analysis of GWAS summary statistics identifies additional variants influencing complex traits. Nat Genet 44, 369–375, S1-3 (2012).

72. Yang, J., Lee, S. H., Goddard, M. E. & Visscher, P. M. GCTA: a tool for genome-wide complex trait analysis. Am J Hum Genet 88, 76–82 (2011).

73. Kichaev, G. et al. Integrating Functional Data to Prioritize Causal Variants in Statistical Fine-Mapping Studies. PLoS Genet 10, e1004722 (2014).

74. Finucane, H. K. et al. Partitioning heritability by functional annotation using genome-wide association summary statistics. Nat. Genet. 47, 1228–1235 (2015).

75. The GTEx Consortium et al. The GTEx Consortium atlas of genetic regulatory effects across human tissues. Science 369, 1318–1330 (2020).

76. Foley, C. N. et al. A fast and efficient colocalization algorithm for identifying shared genetic risk factors across multiple traits. Nat Commun 12, 764 (2021).

77. Barbeira, A. N. et al. Exploring the phenotypic consequences of tissue specific gene expression variation inferred from GWAS summary statistics. Nat Commun 9, 1825 (2018).

78. Barbeira, A. N. et al. Integrating predicted transcriptome from multiple tissues improves association detection. PLoS Genet 15, e1007889 (2019).

79. Gusev, A. et al. Integrative approaches for large-scale transcriptome-wide association studies. Nat Genet 48, 245–252 (2016).

80. Zhang, J. et al. Plasma proteome analyses in individuals of European and African ancestry identify cis-pQTLs and models for proteome-wide association studies. Nat Genet 54, 593–602 (2022).

81. Zhu, Z. et al. Integration of summary data from GWAS and eQTL studies predicts complex trait gene targets. Nat Genet 48, 481–487 (2016).

82. Min, J. L. et al. Genomic and phenotypic insights from an atlas of genetic effects on DNA methylation. Nat Genet 53, 1311–1321 (2021).

83. Relton, C. L. et al. Data Resource Profile: Accessible Resource for Integrated Epigenomic Studies (ARIES). Int. J. Epidemiol. 44, 1181–1190 (2015).

84. Hatton, A. A. et al. Genetic control of DNA methylation is largely shared across European and East Asian populations. Nat Commun 15, 2713 (2024).

85. McRae, A. F. et al. Identification of 55,000 Replicated DNA Methylation QTL. Sci Rep 8, 17605 (2018).

86. Hannon, E. et al. Leveraging DNA-Methylation Quantitative-Trait Loci to Characterize the Relationship between Methylomic Variation, Gene Expression, and Complex Traits. Am J Hum Genet 103, 654–665 (2018).

87. Hannon, E. et al. An integrated genetic-epigenetic analysis of schizophrenia: evidence for co-localization of genetic associations and differential DNA methylation. Genome Biol 17, 176 (2016).

88. Mortlock, S. et al. Global endometrial DNA methylation analysis reveals insights into mQTL regulation and associated endometriosis disease risk and endometrial function. Commun Biol 6, 780 (2023).

89. de Leeuw, C. A., Mooij, J. M., Heskes, T. & Posthuma, D. MAGMA: generalized gene-set analysis of GWAS data. PLoS Comput Biol 11, e1004219 (2015).

90. Frei, O. et al. Improved functional mapping of complex trait heritability with GSA-MiXeR implicates biologically specific gene sets. Nat Genet 56, 1310–1318 (2024).

91. Ge, T., Chen, C.-Y., Ni, Y., Feng, Y.-C. A. & Smoller, J. W. Polygenic prediction via Bayesian regression and continuous shrinkage priors. Nat Commun 10, 1776 (2019).

92. Hemani, G. et al. The MR-Base platform supports systematic causal inference across the human phenome. eLife 7, e34408 (2018).

93. Pierce, B. L. & Burgess, S. Efficient design for Mendelian randomization studies: subsample and 2-sample instrumental variable estimators. Am J Epidemiol 178, 1177– 1184 (2013).

94. Bowden, J., Davey Smith, G., Haycock, P. C. & Burgess, S. Consistent Estimation in Mendelian Randomization with Some Invalid Instruments Using a Weighted Median Estimator. Genet Epidemiol 40, 304–314 (2016).

95. Hartwig, F. P., Davey Smith, G. & Bowden, J. Robust inference in summary data Mendelian randomization via the zero modal pleiotropy assumption. Int J Epidemiol 46, 1985–1998 (2017).

96. Verbanck, M., Chen, C.-Y., Neale, B. & Do, R. Detection of widespread horizontal pleiotropy in causal relationships inferred from Mendelian randomization between complex traits and diseases. Nat Genet 50, 693–698 (2018).

97. Hemani, G., Tilling, K. & Davey Smith, G. Orienting the causal relationship between imprecisely measured traits using GWAS summary data. PLoS Genet 13, e1007081 (2017).

98. Bowden, J. et al. Assessing the suitability of summary data for two-sample Mendelian randomization analyses using MR-Egger regression: the role of the I2 statistic. Int J Epidemiol 45, 1961–1974 (2016).

99. Burgess, S. & Thompson, S. G. Interpreting findings from Mendelian randomization using the MR-Egger method. Eur J Epidemiol 32, 377–389 (2017).

